# Suicide notes as structured final communications: evidence from 100,000 deaths in South Korea

**DOI:** 10.64898/2026.10.05.26364805

**Authors:** Jae Bin Park, Adedoyin Aderinwale, Seung Yoon Oh, Young Hye Hur, Soon Ju Lee, Se Won Kwon, Jaeseung Jeong

**Affiliations:** Department of Brain and Cognitive Sciences, Korea Advanced Institute of Science and Technology, Daejeon, South Korea; Korea Foundation for Suicide Prevention, Research Evaluation Department; Korea Foundation for Suicide Prevention, Counselling Support Team; Korea Foundation for Suicide Prevention, Policy Research Department

**Keywords:** nationwide suicide data, Korea, suicide notes, demographic comparison, section-based analysis, sentiment analysis, thematic analysis

## Abstract

Humans often communicate meaningfully during existential transition, yet the structure of final communications before death remains poorly understood. Suicide notes provide rare behavioral records, but two questions remain unresolved: whether note-leavers represent the broader population of suicide decedents, and whether notes follow systematic narrative structures. We analyzed a nationwide police-investigated registry of 102,922 suicide deaths in South Korea from 2013 to 2020, including 38,044 notes. Note-leavers differed systematically from non-leavers across demographic, situational and behavioral characteristics, indicating that note production reflects a selective communicative act within the suicidal process. Modeling notes as sequential narratives, we identified consistent patterns across opening, middle and closing sections, organized around apology and shame, love and gratitude, and practical directives concerning post-mortem affairs. Emotional trajectories also varied by age and gender. These findings provide population-scale evidence that suicide notes are structured final communications shaped by social relationships and demographic context under extreme circumstances.

## INTRODUCTION

Human beings often communicate meaningfully at moments of existential transition. Final messages left before death appear across cultures and historical periods, from soldiers’ farewell letters to last statements delivered before execution. These communications are rarely random expressions of emotion; rather, they frequently serve social, moral, and practical purposes such as repairing relationships, expressing gratitude, assigning responsibility, or organizing matters left unresolved. Despite the apparent universality of such behaviors, the structure and determinants of end-of-life communication remain poorly understood.

Suicide notes provide one of the few direct behavioral records of cognition immediately preceding death. As such, they offer a rare opportunity to observe how individuals interpret, justify, and communicate their final decisions. For over a century, suicide has been understood not only as an individual psychological event but also as a social phenomenon shaped by relationships and collective norms ^1^, and the complex interplay between the psychological, social, and environmental factors involved remains poorly understood ^2^. Suicide notes therefore represent an important interface between individual psychology and social meaning, reflecting how individuals position their actions within a network of relationships and responsibilities, addressing family members, friends, and society at large ^3^ and presents possibilities for detecting suicide risk using text ^4^.

Accordingly, suicide notes have long been studied as psychological artifacts of suicidal cognition ^3,5^. Early qualitative analyses identified recurring thematic elements such as apology, affection toward loved ones, explanations of distress, and instructions concerning post-mortem affairs ^5–7^. Subsequent linguistic and computational analyses demonstrated that suicide notes contain distinctive emotional and semantic patterns that differentiate genuine notes from simulated ones and reveal systematic patterns of affective expression ^8,9^. More recent computational approaches have further shown that suicide notes exhibit structured semantic organization and cognitive network properties ^10^. These patterns often reflect psychological constructs central to theories of suicidal behavior, including perceived burdensomeness, hopelessness, and interpersonal attachment ^11–13^. The interpersonal theory identifies perceived burdensomeness and thwarted belongingness as central drivers of suicidal desire. ^11,13^. Suicide notes frequently contain language consistent with these constructs, suggesting that they provide behavioral traces of the social and cognitive processes underlying suicidal ideation.

Despite these insights, empirical research on suicide notes remains constrained by important limitations. Most studies rely on relatively small samples, often consisting of only a few hundred notes, limiting the ability to robustly examine how note content varies across demographic and situational contexts ^14,15^. Additionally, many analyses treat suicide notes as undifferentiated textual documents, examining word frequencies or emotional categories across entire texts rather than the narrative progression of how content unfolds sequentially across a message.

A further unresolved question concerns the representativeness of individuals who left suicide notes (hereafter, note-leavers). Not all individuals who die by suicide leave written messages, and understanding who leaves suicide notes is essential for interpreting note-based research. Writing a suicide note requires both the intention to communicate and the opportunity to do so, making note production a potentially selective behavioral act within the suicidal process. Early studies reported relatively few differences between note-leavers and non-leavers ^16,17^. However, more recent research suggests that note-leavers may differ systematically from non-leavers across demographic, clinical, and situational characteristics, including age, mental health history, and suicide method ^18,19^. If suicide note-leavers represent a distinct subgroup, conclusions drawn from suicide note content may not generalize to the broader population of suicide decedents.

Another largely unexplored question concerns the internal organization of suicide notes themselves. Written communication rarely occurs as an unstructured stream of emotion. Instead, messages typically follow recognizable narrative architectures in which authors introduce relational context, articulate emotional states or explanations, and conclude with expressions of closure or practical instruction. Although prior research has identified common thematic elements in suicide notes ^5,7^, little is known about how these themes are arranged across the progression of a message. Investigating this narrative structure may therefore provide insight into how individuals organize meaning and social relationships when composing their final communications.

Demographic and sociocultural factors may further shape these communicative patterns. Suicide is embedded within networks of social relationships and cultural expectations concerning responsibility, honor, and interpersonal obligation ^1^. Age and gender represent particularly important dimensions of variation. Life-course perspectives suggest that individuals at different stages of life may prioritize different communicative goals. Younger individuals may emphasize emotional expression and interpersonal reconciliation, whereas older individuals may focus more on practical closure and responsibility toward surviving relatives ^6,20^. Gender differences in emotional expression and social roles may likewise influence how individuals frame their final messages ^14,21,22^. However, large-scale empirical evidence examining how demographic factors shape the narrative structure of suicide notes remains limited.

South Korea provides an important context in which to examine these questions as it consistently reports one of the highest suicide rates among developed countries and reflects a complex interplay of socioeconomic pressures, rapid social change, and evolving family structures ^23^. South Korean cultural norms influenced by Confucian traditions emphasize social obligation, relational harmony, and the avoidance of shame, potentially shaping how individuals frame responsibility and apology in final communications ^24^.

In the present study, we integrate demographic, situational, and linguistic analyses using a nationwide registry of police-investigated suicides in South Korea spanning eight years (2013–2020). The dataset includes 102,922 suicide cases, of which more than 38,000 contain suicide notes, representing one of the largest collections of suicide notes analyzed to date. Leveraging this population-scale dataset, we address three questions central to understanding suicide notes as final communications. First, we examine who leaves suicide notes by identifying demographic, situational, and psychosocial characteristics that distinguish note-leavers from non-leavers. Second, we investigate how suicide notes are internally organized by modeling them as sequential narratives consisting of opening, middle, and closing sections. Third, we examine how thematic and emotional trajectories within these narratives vary across age and gender.

By integrating population-scale behavioral data with computational linguistic analysis, we demonstrate that suicide notes are not merely expressions of distress but structured final communications shaped by demographic context, interpersonal relationships, and communicative goals. From a broader perspective, our findings suggest that end-of-life written messages constitute an organized form of human social communication, providing a framework for understanding how individuals negotiate meaning, responsibility, and social relationships under extreme psychological circumstances.

## RESULTS

### Correlates of note leaving

#### National counts and temporal patterning

The registry comprised 102,922 suicides between 2013 and 2020, of which 38,044 (36.9%) indicated the presence of a suicide note. Absolute counts were highest in Gyeonggi and Seoul, whereas population-normalized annual rates were highest in Gangwon and South Chungcheong (Fig. 1a). Monthly counts of note-leavers and non-leavers were strongly correlated (Pearson’s r = 0.78, p < 0.001), indicating shared seasonal variation in overall suicide incidence. However, the proportion of suicides with notes increased toward the end of the calendar year (Fig. 1b), suggesting that note leaving showed additional within-year variation beyond the seasonality of suicide counts.

**Figure 1.**
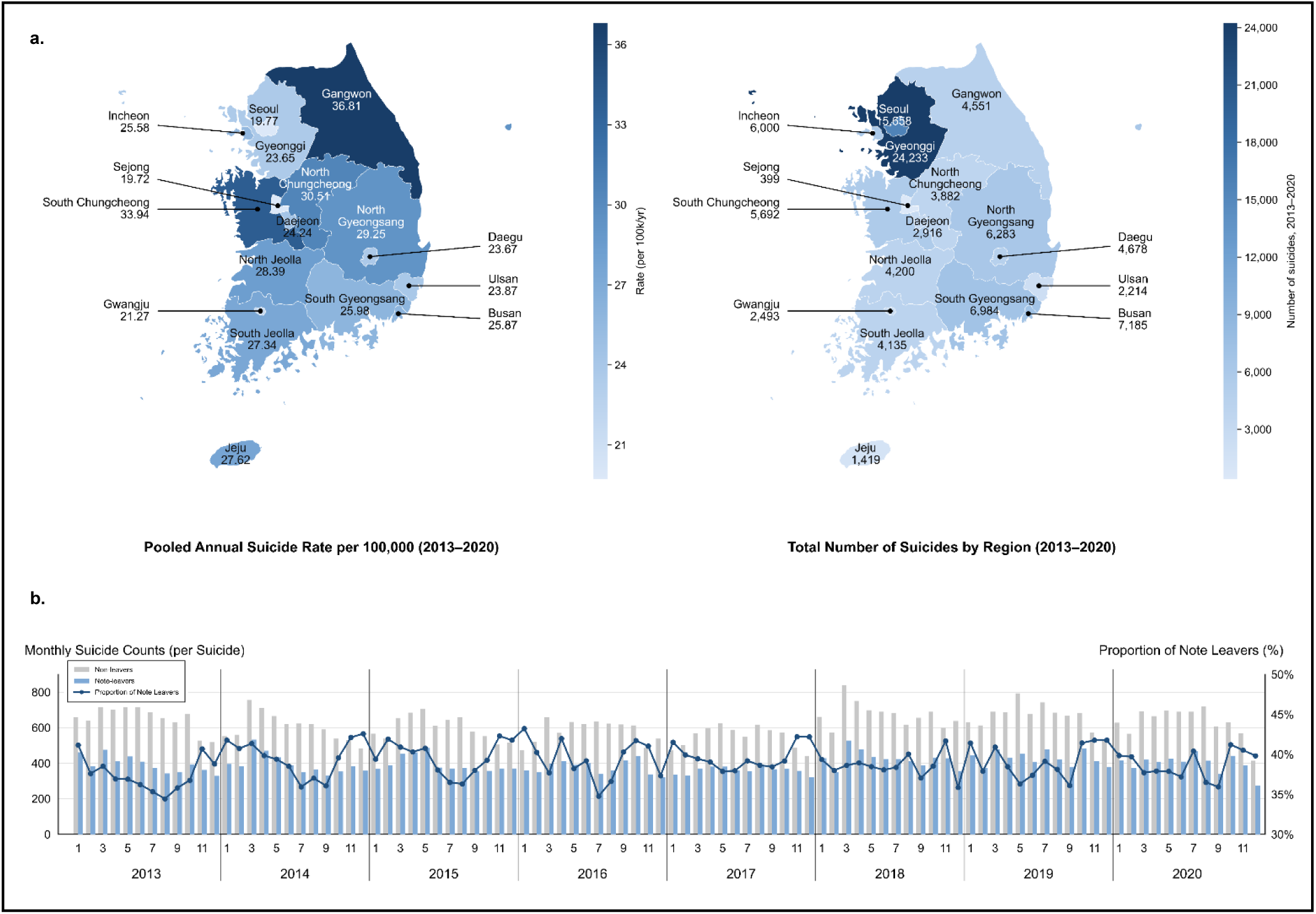
Regional suicide counts and temporal dynamics of suicide across note-leaving in South Korea (2013–2020). (a) Pooled annual suicide rate per 100,000 by region, shaded by Σ yearly deaths ÷ Σ yearly mid-year population × 100,000 (darker shading = higher rates) (Left) and total suicides counts by region, 2013–2020 (darker shading = higher counts) (Right). (b) Monthly suicide counts stratified by note-leavers (blue) and non-leavers (gray); the series are strongly correlated (Pearson r = 0.78, p < 0.001). Monthly proportion of note-leavers, showing recurrent late-year peaks. Population denominators from KOSIS (Korean Statistical Information Service).

#### Sociodemographic and situational correlates

We next estimated odds ratios (ORs) from logistic regression models predicting note leaving from demographic and situational covariates (reference categories in Methods; Fig. 2). Effects were generally modest in magnitude but consistent across domains.

**Figure 2.**
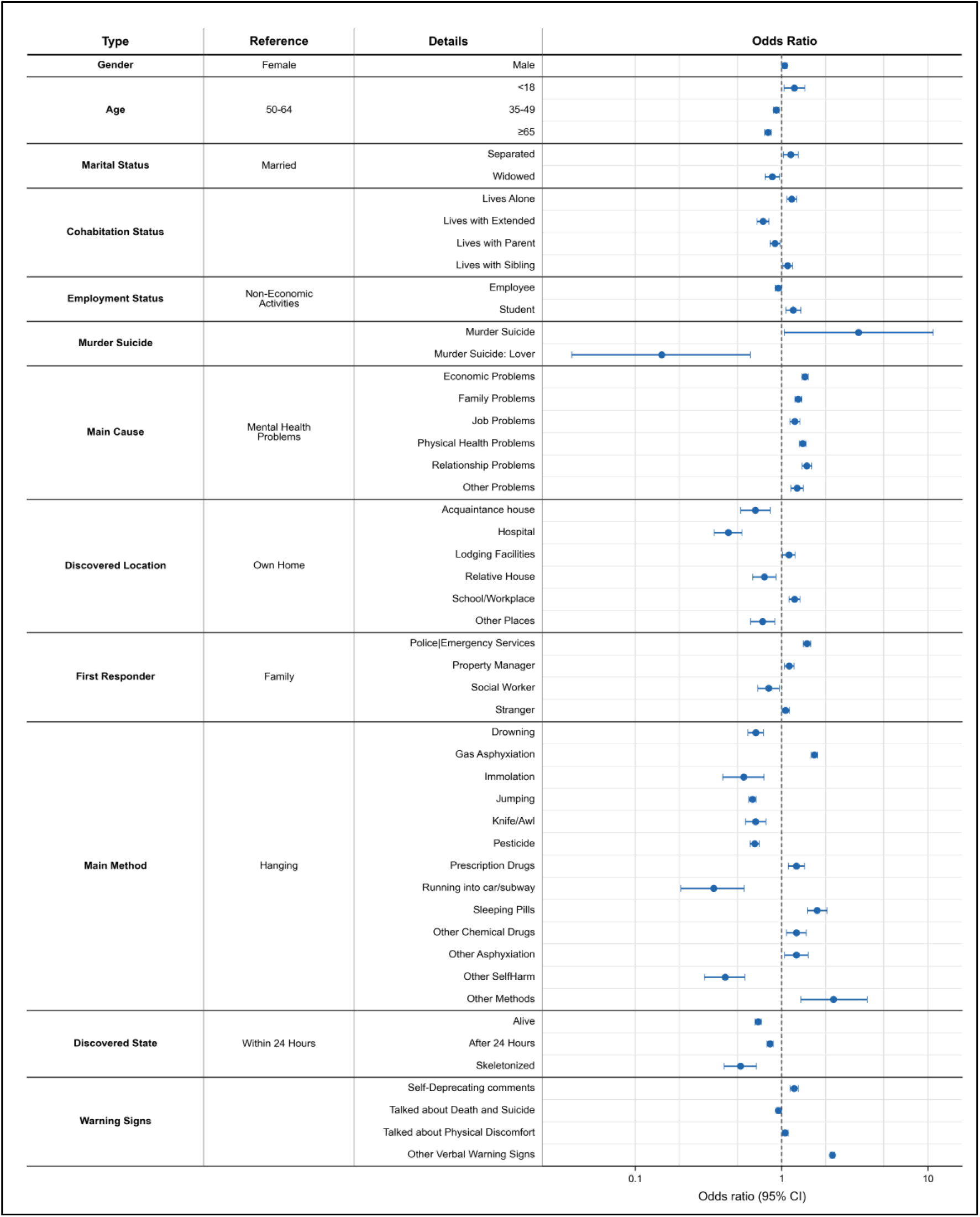
Multivariate logistic regression derived odds ratios of leaving suicide notes across statistically significant variables –. Adjusted odds ratio across statistically significant variables are presented. Odds ratio scores that are higher than 1 represents greater odds of leaving a note while odds ratio scores lower than 1 represents greater odds of not leaving a note.

Note leaving was slightly more common among males (OR = 1.05, 95% CI 1.00–1.09) and those aged ≤18 years (OR = 1.22, 95% CI 1.04–1.44), and less common among those aged 35–49 years (OR = 0.92, 95% CI 0.88–0.96) and ≥65 years (OR = 0.80, 95% CI 0.76–0.85), relative to the reference age group. Relative to married individuals, note leaving was more common among those who were separated (OR = 1.15, 95% CI 1.02–1.29) or living alone (OR = 1.17, 95% CI 1.09–1.26), and less common among widowed individuals (OR = 0.86, 95% CI 0.77–0.96).

Across cohabitation categories, note leaving was more common among those living with siblings (OR = 1.10, 95% CI 1.02–1.18) and less common among those living with parents (OR = 0.90, 95% CI 0.83–0.97) or other relatives (OR = 0.74, 95% CI 0.68–0.82). Socioeconomic differences were smaller: relative to non-economic activity, note leaving was more common among students (OR = 1.20, 95% CI 1.07–1.35) and slightly less common among employees (OR = 0.95, 95% CI 0.90–0.99), with no reliable differences across occupational categories.

#### Circumstances, method and pre-death communication

Circumstantial factors also differentiated note-leavers from non-leavers. Cases involving economic, physical-health, family, relationship, and job-related problems were more likely to include a note (ORs = 1.23–1.48), and murder–suicide cases also showed elevated odds (OR = 3.35, 95% CI 1.04–10.80). Compared with suicides occurring at home, note leaving was more common in lodging facilities (OR = 1.12, 95% CI 1.02–1.23) and at school or work (OR = 1.22, 95% CI 1.12–1.33), and less common in hospitals (OR = 0.43, 95% CI 0.35–0.53) or at relatives’ and friends’ homes (OR = 0.76 and 0.66). Discovery was more often by strangers, police or emergency personnel, and property managers (OR = 1.06, 1.49, and 1.12), whereas social workers more often discovered non-leavers (OR = 0.81, 95% CI 0.69–0.96). Time-of-day distributions did not differ.

Suicide method also varied systematically. Relative to hanging, note leaving was more common in gas asphyxiation (OR = 1.67, 95% CI 1.59–1.76), prescribed drugs (OR = 1.26, 95% CI 1.11–1.43), sleeping pills (OR = 1.75, 95% CI 1.50–2.04), and other chemicals or asphyxiants (OR = 1.26, 95% CI 1.04–1.52), and less common in jumping (OR = 0.63, 95% CI 0.60–0.67), self-immolation (OR = 0.55, 95% CI 0.40–0.76), traffic or subway jumping (OR = 0.34, 95% CI 0.20–0.55), pesticide (OR = 0.65, 95% CI 0.61–0.70), drowning (OR = 0.66, 95% CI 0.59–0.75), and stabbing (OR = 0.66, 95% CI 0.57–0.78). Note-leavers were also less likely to be found alive (OR = 0.69, 95% CI 0.66–0.72), discovered after 24 hours (OR = 0.83, 95% CI 0.79–0.87), or found in a skeletonized state (OR = 0.52, 95% CI 0.40–0.67).

Reported verbal cues before death also differed. Note-leavers more often expressed physical discomfort (OR = 1.06, 95% CI 1.01–1.11), self-deprecating comments (OR = 1.22, 95% CI 1.14–1.30), and nonspecific distress (OR = 2.22, 95% CI 2.14–2.30), whereas explicit statements about suicide or death were slightly less common (OR = 0.95, 95% CI 0.92–0.98) (Fig. 2). Taken together, these findings indicate that note leaving was associated with reproducible differences in demographic profile, social context, method, and pre-death communication.

#### Lexical profile and thematic clustering

To characterize the broad lexical profile of notes, we examined the most frequent noun and verb categories across all notes. Kinship and family terms were the most frequent noun category (14.63%), and apology-related verbs were the most frequent verb category (13.63%) (Fig. 3), indicating that interpersonal ties and contrition were central features of final written communications.

**Figure 3.**
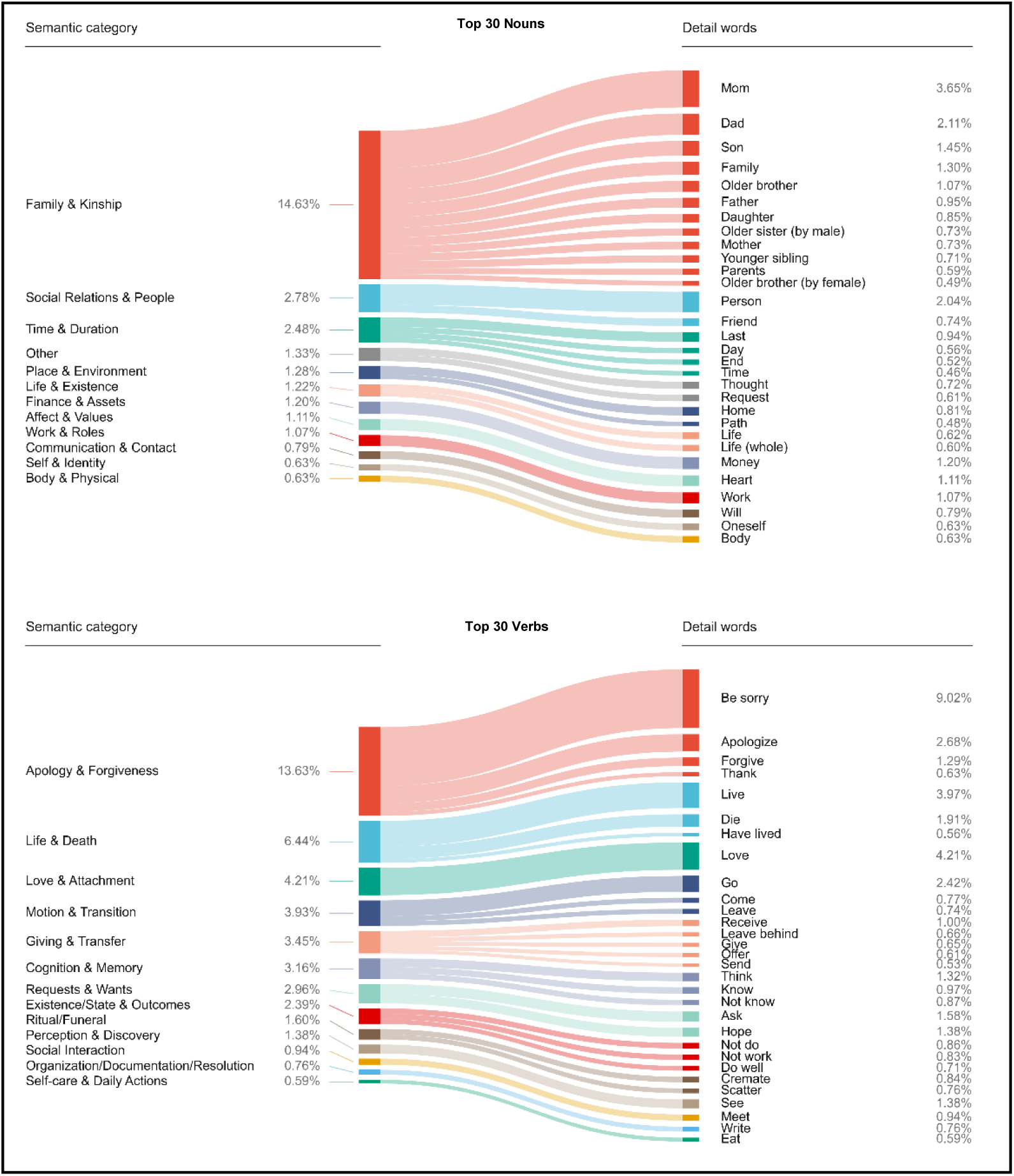
Translated noun and verb frequencies based on suicide notes –. Fig 3 displays the most frequently occurring nouns (3a) and verbs (3b) in suicide notes from the dataset. The Inner bar plot represents the related topics of words and the percentage of topics across the top 50 nouns and verbs in the notes. Nouns related with family and kinship, and verbs related with apology and forgiveness were dominant.

We next examined whether notes showed recurring thematic organization when modeled as sectioned communications. In PCA space, theme clusters showed clear separation (cumulative explained variance = 0.53; silhouette score = 0.49; Fig. 4a and Extended Data 2a). Cluster profiles were organized primarily around Sorry/Shame, Love/Gratitude, and Post-mortem Affairs. Sorry/Shame frequently appeared as the dominant theme within a single section, recurred across two sections, or extended across all three. Love/Gratitude commonly appeared either as a sustained theme or in combination with Sorry/Shame in early sections. Post-mortem Affairs was concentrated later in notes, most often in middle-to-ending trajectories. By contrast, Burden and Despair rarely dominated cluster centroids and, when present, were typically confined to the middle section.

**Figure 4.**
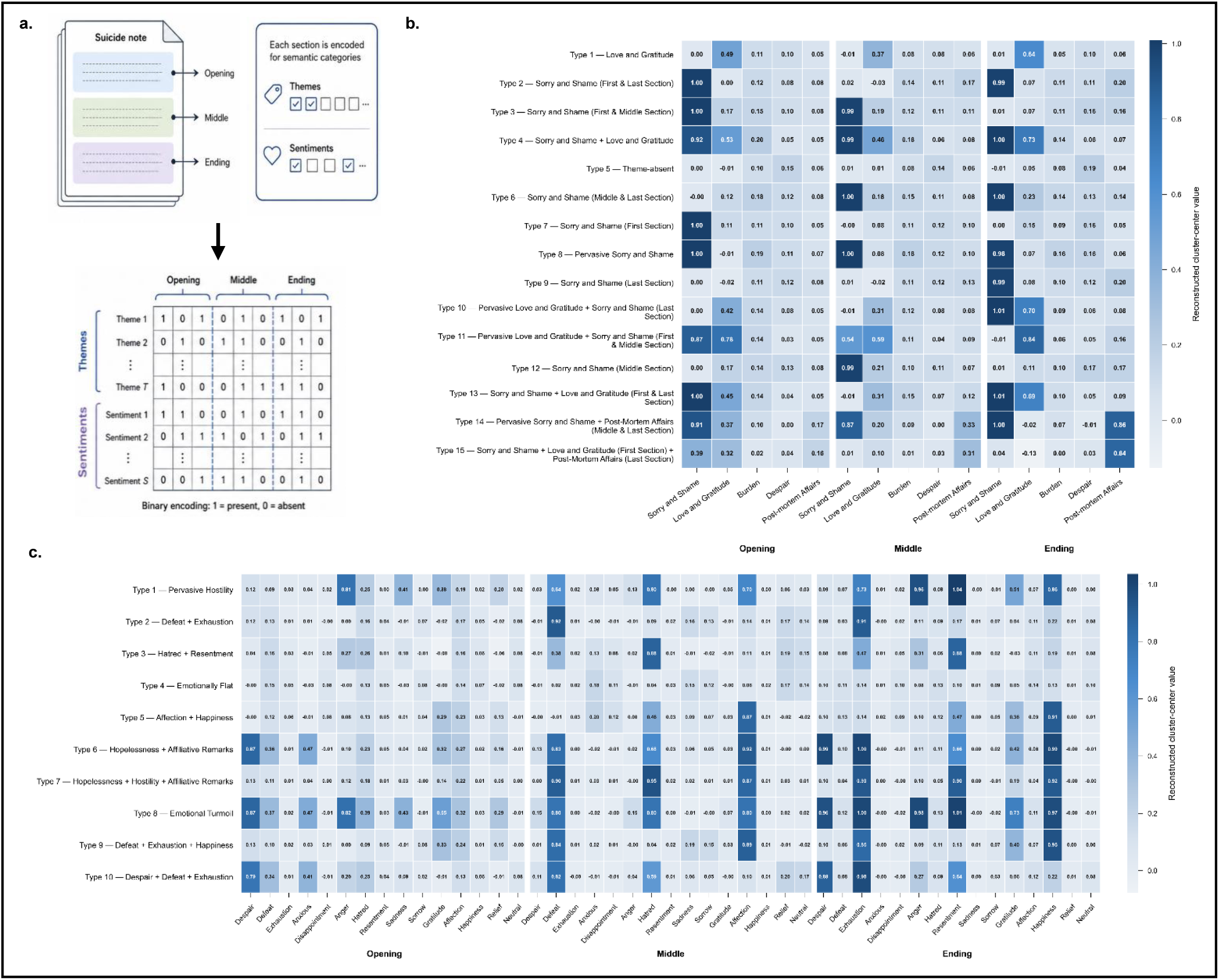
Vectorizing suicide notes and PCA-reduced K-means Clusters of Theme and Sentiment Matrices –. Fig 4a. presents a summary visualization of the suicide notes and how they are preprocessed for the current study. The image displays how suicide notes are first split into introduction, body and conclusion and vectorized based on the presence of specific themes or sentiments in the suicide notes. Fig 4b. and Fig 4c. show K-means clusters derived from theme matrices and sentiment matrices with labels across each cluster. Based on the explained variance and Silhouette score, a total of 15 theme clusters and 10 sentiment were used for clustering. Explanation of each clusters are provided in Supplementary Table 3.

#### Theme placement, transitions and demographic patterning

We then quantified both where themes occurred and how they progressed across sections using transition analyses and section-specific logistic regression models (Fig. 5). Across notes, transitions tended to converge on Sorry/Shame, consistent with its high overall prevalence. Gender differences were most evident in early thematic sequencing: from opening to middle sections, transitions between Love/Gratitude and Sorry/Shame differed by gender (ΔLift = −0.14, −0.23, and −0.10; permutation *p* < 0.05), and Post-mortem Affairs also showed significant persistence (*p* < 0.01). Age differences were more pronounced among older adults (≥65 years). From opening to middle sections, Burden showed enriched persistence (Burden→Burden: ΔLift = 0.62, *p* < 0.001) and increased transitions into Post-mortem Affairs (Burden→Post-mortem Affairs: ΔLift = 0.40, *p* < 0.01) (Fig. 7a, c, e).

**Figure 5.**
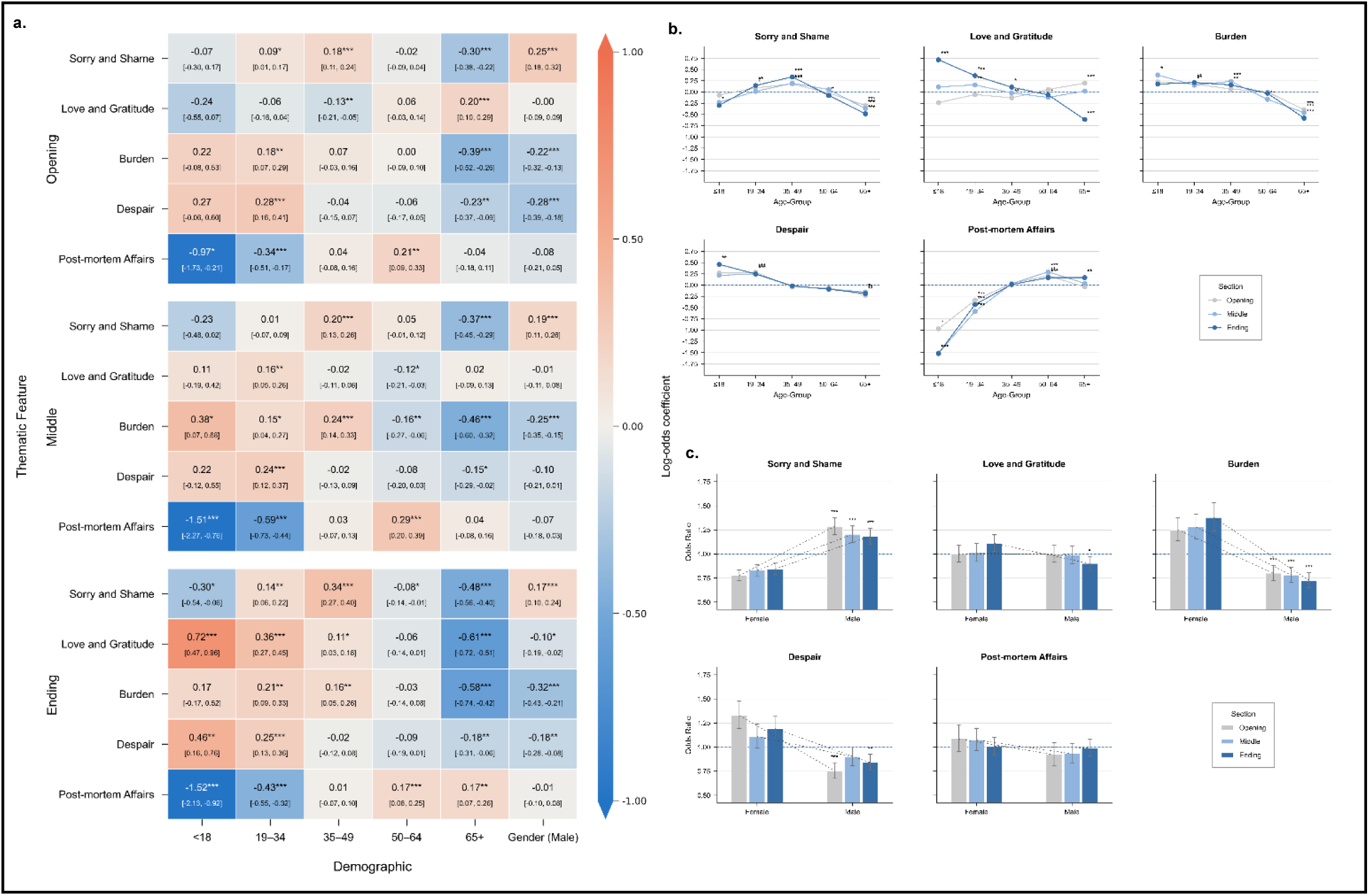
Theme-based logistic regression across age groups and gender –. Fig 5a. presents the log odds ratios (with CIs) across age and gender. The log odds ratio are presented across sections and significances are labelled based on FDR-adjusted p-values, with *FDR-adjusted p < 0.05, **FDR-adjusted p < 0.01 and ***FDR-adjusted p < 0.001. Fig 5b. presents the theme-based log odds ratios across age groups and Fig 5c. presents the theme-based log odds ratios across gender in all themes and sections.

Section-specific models showed parallel age gradients in thematic prevalence. Burden and Despair declined with age across sections, with the lowest prevalence in the ≥65 group (for example, Burden ending log OR = −0.58, FDR-adjusted p < 0.001; Despair ending log OR = −0.18, FDR-adjusted p < 0.01). Sorry/Shame followed an inverse U-shaped age pattern, peaking in ages 35–49 (ending log OR = 0.34, FDR-adjusted p < 0.001) and appearing less often among both the youngest (≤18 ending log OR = −0.30, FDR-adjusted p < 0.05) and oldest groups (≥65 ending log OR = −0.48, FDR-adjusted p < 0.001). Love/Gratitude was elevated in ending sections among younger groups (≤18 log OR = 0.72, FDR-adjusted p < 0.001; 19–34 log OR = 0.36, FDR-adjusted p < 0.001), but in opening sections among older adults (≥65 log OR = 0.20, FDR-adjusted p < 0.001). Post-mortem Affairs was sharply reduced in younger groups (≤18 middle log OR = −1.51, ending log OR = −1.52; both FDR-adjusted p < 0.001; 19–34 middle log OR = −0.59, FDR-adjusted p < 0.001), but more common in midlife (50–64 middle log OR = 0.29, FDR-adjusted p < 0.001) and in ending sections among older adults (≥65 ending log OR = 0.17, FDR-adjusted p < 0.01).

Gender effects were also reliable after FDR correction. In the model coding used, Sorry/Shame was more prevalent in male-authored notes across sections (opening/middle/ending log ORs = 0.25, 0.19, and 0.17; all FDR-adjusted p < 0.001). Additional FDR-significant gender effects were observed for Burden across sections, Despair in the opening and ending sections, and Love/Gratitude in the ending section (Fig. 5), whereas Post-mortem Affairs showed minimal gender differences (all |log OR| ≤ 0.08, all FDR-adjusted p > 0.25). Overall, these findings indicate that demographic variation in suicide-note themes was expressed not only in whether themes appeared, but also in where they appeared and how they developed across sections.

#### Emotional organization across note sections

We applied the same approach to section-level sentiment matrices. Sentiment clusters showed weaker separation than thematic clusters (cumulative explained variance = 0.51; silhouette score = 0.22; Fig. 4b and Extended Data 2b), but still revealed recurring profiles. These were dominated by negative affective states, including hostility-related profiles, defeat/exhaustion, hatred/resentment, and broader mixed distress patterns that often persisted or intensified toward the ending section. Two mixed profiles combined strong hopelessness with affiliative sentiment, indicating that affiliative language often co-occurred with negative states rather than replacing them. Positive profiles were less common, and relief was rare and tended to occur late.

#### Sentiment placement, transitions and demographic patterning

We next examined section-to-section sentiment transitions and section-specific demographic associations (Fig. 6). Gender differences were most pronounced in early and late affective sequencing. From opening to middle sections, female-authored notes showed greater persistence within negative affective states, including sadness, disappointment, and despair (Sadness→Sadness: ΔLift = −0.04, *p* < 0.001). Male-authored notes showed relatively greater redirection from self-evaluative negative states toward higher-arousal antagonistic affect. From middle to ending sections, male-authored notes showed relatively more transitions from anxiety and exhaustion into despair and sorrow, whereas female-authored notes showed comparatively fewer re-entries into high-intensity distress pathways.

**Figure 6.**
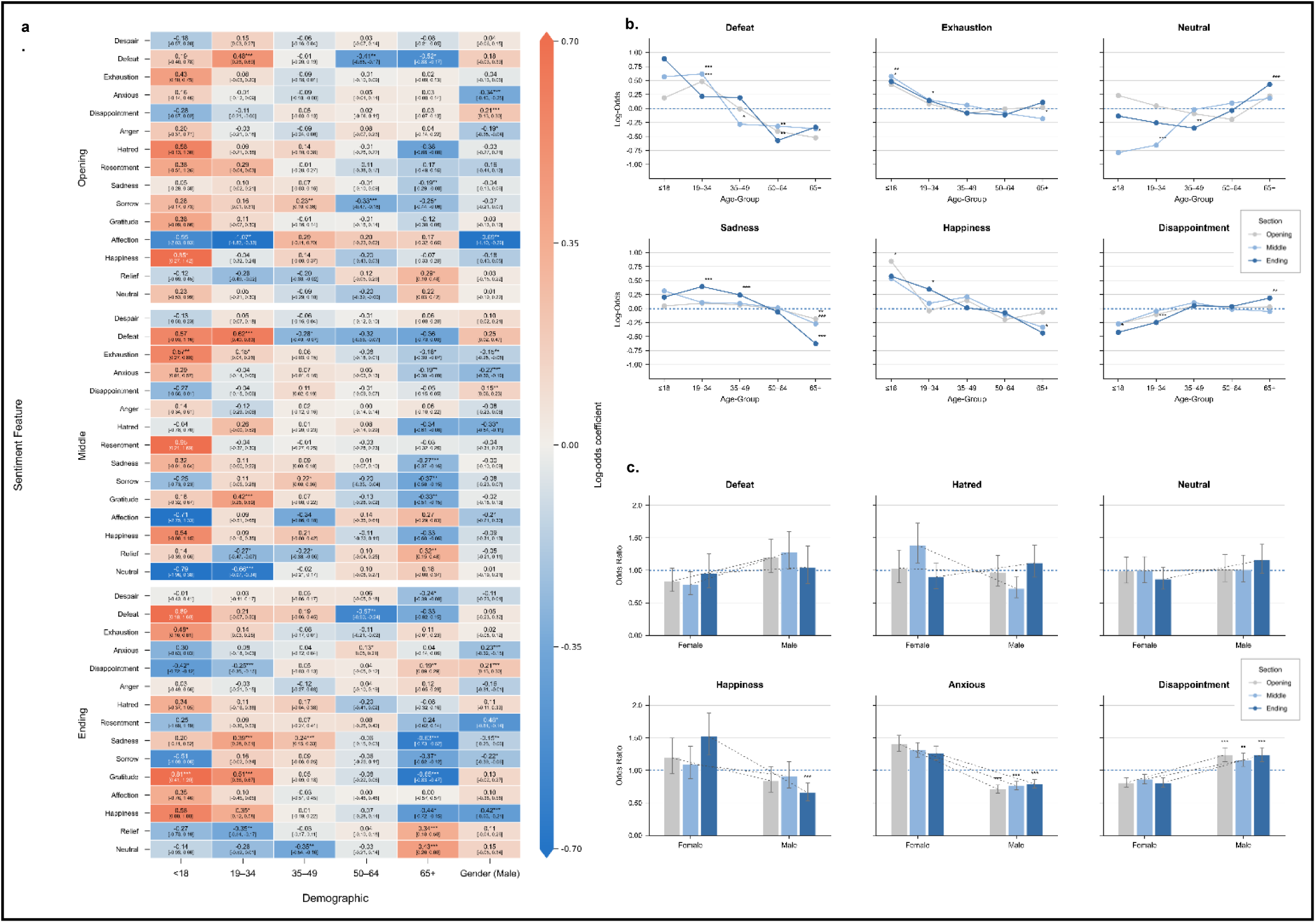
Sentiment-based logistic regression across age groups and gender -. **Fig 6a**. presents the log odds ratio (with CIs) across age and gender. The log odds ratios are presented across sections and significances are labelled based on FDR-adjusted p-values, with *FDR-adjusted p < 0.05, **FDR-adjusted p < 0.01 and ***FDR-adjusted p < 0.001. Fig 6b. presents the theme-based log odds ratio across age groups and Fig 6c. presents the theme-based log odds ratio across gender in all themes and sections.

Age differences were again most pronounced among older adults. From opening to middle sections, Defeat showed strong persistence (Defeat→Defeat: ΔLift = 7.22, *p* < 0.001) and enriched transitions into Despair and Exhaustion (ΔLift = 1.07 and 1.04; both *p* < 0.001). Older-adult notes also showed enriched transitions from distress into Gratitude and Relief (ΔLift = 0.30 and 0.31; both *p* < 0.01). From middle to ending sections, older-adult notes showed enriched transitions into Sorrow (Despair→Sorrow: ΔLift = 0.98, *p* < 0.001), strong persistence of Sorrow (Sorrow→Sorrow: ΔLift = 1.45, *p* < 0.001), and reduced persistence of Neutral (Neutral→Neutral: ΔLift = −2.80, *p* < 0.01) (Fig. 7b, d, f).

**Figure 7.**
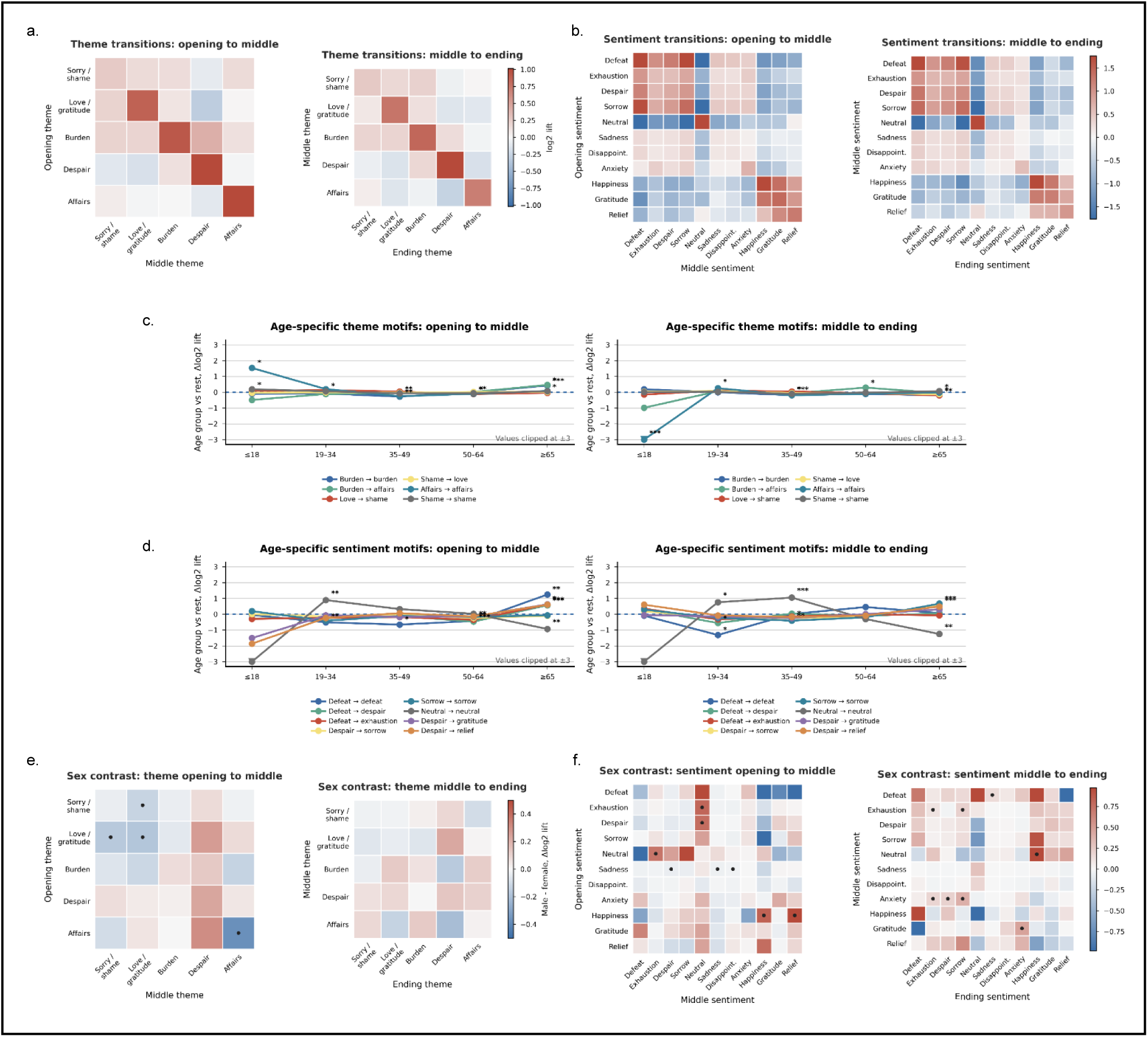
Theme and sentiment transitions across ordered sections of suicide notes. (**a,b)** Overall section-to-section transition enrichment for themes (a) and sentiments (b) from opening to middle and from middle to ending sections. Heatmap colors show log₂ lift, where positive values indicate transitions occurring more often than expected from the later-section base prevalence. **(c,d)** Age-specific transition motifs for themes (c) and sentiments (d), expressed as age-group-versus-rest Δlog₂ lift. Motifs were selected a priori from the principal results, including burden persistence, burden-to-affairs transitions, love/gratitude–sorry/shame transitions, defeat persistence, transitions into despair, exhaustion and sorrow, neutral persistence and transitions from distress into gratitude or relief. **(e,f)** Sex differences in transition enrichment for themes (d) and sentiments (f) across both adjacent section intervals. Values show male–female Δlog₂ lift; positive values indicate enrichment in male-authored notes and negative values indicate enrichment in female-authored notes. Black dots indicate cellwise permutation *P* < 0.05. Overall, the figure shows that demographic differences in suicide-note content were expressed not only in the prevalence of specific themes and sentiments, but also in their sequential placement and redirection across the note.

Section-specific models showed consistent age gradients in emotional prevalence. Defeat and Exhaustion declined with age (Defeat opening 65+ log OR = −0.52, FDR-adjusted p < 0.05; Exhaustion middle 65+ log OR = −0.18, FDR-adjusted p < 0.05). Happiness also declined with age (65+ ending log OR = −0.44, FDR-adjusted p < 0.05), whereas Neutral was more prevalent in ending sections among older adults (65+ log OR = 0.43, FDR-adjusted p < 0.001). Disappointment increased with age (65+ ending log OR = 0.19, FDR-adjusted p < 0.01). Sadness peaked in young adults (19–34 ending log OR = 0.39, FDR-adjusted p < 0.001) and was reduced in late life (65+ ending log OR = −0.63, FDR-adjusted p < 0.001).

Gender effects were concentrated in evaluative and higher-arousal emotions. In the model coding used, male-authored notes showed a positive middle-section estimate for Defeat (log OR = 0.25), but this association did not survive FDR correction (FDR-adjusted p = 0.090). Male-authored notes showed higher Disappointment across sections (opening and ending FDR-adjusted p < 0.001; middle FDR-adjusted p < 0.01). Female-authored notes showed higher Anxiety across sections (all FDR-adjusted p < 0.001) and higher Happiness in the ending section (log OR = −0.42, FDR-adjusted p < 0.001). Additional FDR-significant differences were observed for Hatred in the middle section (FDR-adjusted p = 0.013), Anger in the opening section (FDR-adjusted p = 0.038), and Affection in the opening section (FDR-adjusted p = 0.005). Gender differences in Neutral were small and not significant after correction (all FDR-adjusted p ≥ 0.289). Together, these analyses show that demographic differences in suicide-note affect were reflected not only in the prevalence of particular sentiments but also in their placement, persistence, and redirection across the note.

## DISCUSSION

Using a nationwide registry of 102,922 police-investigated suicides in South Korea, we examined two linked questions: whether suicide note-leavers differ systematically from non-leavers, and whether suicide notes show structured internal organization when analyzed as sequential communications rather than as undifferentiated texts. Across analyses, two conclusions emerged. First, suicide note leaving was not randomly distributed across suicide decedents, but was associated with consistent demographic, situational, behavioral and communicative correlates. Second, suicide notes were not homogeneous expressions of distress: thematic and emotional content showed non-random placement and sequencing across opening, middle and closing sections, with additional variation by age and gender. Together, these findings suggest that suicide notes are best understood as selective and internally organized behavioral traces, produced by a non-random subset of suicide decedents and structured through recurring communicative patterns that vary by demographic and social context.

These findings address a broader inferential problem in behavioral science. Naturalistic texts are often treated as windows into underlying psychological states, yet their interpretation depends on two conditions that are frequently overlooked: who produces the text, and how the text is organized once produced. Suicide notes provide an unusually consequential case of this problem. If note-leavers differ from non-leavers, note-based findings cannot automatically be generalized to all suicide decedents. If notes are internally organized communications, analyses that collapse them into whole-document summaries may discard behaviorally meaningful information. Inference from final written communications therefore requires attention to both selection into expression and structure within expression.

Our findings reinforce the view that suicide note leaving is not random. Note-leavers differed from non-leavers across age, sex, marital and cohabitation status, suicide method, location, discovery context, putative stressors and pre-death verbal warning signs. These patterns are broadly consistent with previous population-based and psychological autopsy studies showing that individuals who leave suicide notes are not fully representative of all suicide decedents ^16–19^. The present study extends this evidence in a substantially larger national sample and shows that note leaving is patterned across multiple domains of the suicidal event, rather than being reducible to a single demographic or clinical characteristic.

Several associations suggest that note leaving is embedded in the circumstances surrounding death rather than reflecting a simple stable trait. Note-leavers were more likely to die in contexts involving social separation, privacy or institutional distance from close kin, including living alone, lodging facilities and school or workplace settings, and were more often discovered by strangers, police or emergency personnel, or property managers. Suicide method also varied systematically, with note leaving more common in deaths involving gas asphyxiation or drug ingestion and less common in several more abrupt or externally visible methods. These patterns should not be interpreted as evidence of motive, but they suggest that note production co-occurs with particular situational constellations involving method, setting, opportunity and interpersonal context. The warning-sign findings further reinforce this point: note-leavers more often showed indirect or low-specificity verbal expressions of distress, whereas explicit statements about suicide or death were slightly less common. Because the registry does not provide detailed information about timing, recipient or interpretation of these communications, these findings cannot establish a pathway toward note writing. They do, however, underscore that note-leavers constitute a behaviorally distinctive subgroup and that note-based findings should be generalized cautiously.

Beyond identifying who leaves suicide notes, our analyses demonstrate that suicide notes possess systematic internal organization. Lexical profiles emphasized kinship, family and apology-related language, and section-level clustering revealed recurring thematic trajectories organized primarily around apology and shame, love and gratitude, and post-mortem affairs. These findings extend earlier qualitative and thematic studies showing that suicide notes frequently contain apology, affection, practical instruction, hopelessness and burden-related content ^3,5–7^. They also align with computational work suggesting that suicide notes contain non-random semantic and emotional organization ^9,10,15^.

The central contribution of the present analysis is not simply that these themes occur, but that they are organized across the progression of the note. Apology and shame emerged as a structurally central theme, often recurring across sections or organizing transitions between them. Love and gratitude frequently appeared in relation to apology and shame, indicating that affiliative language and self-reproach often coexist within the same communicative sequence. Post-mortem affairs were more often concentrated in later sections, suggesting a recurrent movement from relational or emotional expression toward practical closure. By contrast, burden and despair rarely dominated entire cluster profiles and were more often confined to specific portions of the note, particularly middle sections.

This organization matters methodologically. Whole-document analyses can identify which themes or sentiments occur, but they cannot show where they appear or how they unfold. By modeling notes as sequential communications, we show that themes and sentiments follow recurring pathways across sections, revealing information not recoverable from aggregate document-level counts. Love and gratitude, for example, were meaningful not only as themes, but also in their placement within the note; post-mortem affairs were similarly distinguished by their characteristic concentration toward later sections. At the same time, these patterns should be interpreted as features of communication rather than direct readouts of motive or cognition. The observed sequencing of apology, affiliative language, distress and practical directives is consistent with the idea that suicide notes are organized acts of interpersonal communication, but the present data cannot determine authors’ intentions or the psychological functions of particular sequences at the individual level. The strongest claim supported by the findings is therefore structural: suicide-note content is positioned and transitions non-randomly within documents.

Age was a clear axis of variation in both thematic and emotional organization. Younger decedents more often expressed burden-and despair-related themes, as well as defeat, exhaustion and sadness, whereas older adults more often emphasized post-mortem affairs and showed greater prevalence of neutral sentiment and disappointment in later sections. These findings are broadly consistent with prior observations that younger suicide notes more often foreground interpersonal distress and hopelessness, whereas later-life notes more often emphasize practical arrangements, responsibility and closure ^6,25^. They are also compatible with suicide theories highlighting perceived burdensomeness, hopelessness, belongingness and strain as important components of suicidal thinking, while leaving open the possibility that these concerns are expressed differently across the life course ^11–13,26^.

Crucially, age differences were not limited to overall content prevalence. Love and gratitude were more concentrated in ending sections among younger writers but more common in opening sections among older writers, while post-mortem affairs were reduced among the youngest writers and more characteristic of middle-aged and older adults, particularly in later sections. Apology and shame also followed an inverse U-shaped pattern, peaking in middle adulthood. These patterns suggest that life stage shapes the organization of final written communication, although the data do not establish age-related differences in conscious communicative goals. The more defensible conclusion is that age differences in suicide notes involve not only what is expressed, but also where expressions are positioned within the progression of the message.

Gender differences were also evident across both themes and sentiments. Male-authored notes showed higher apology and shame across sections, as well as higher disappointment in selected sections. Female-authored notes showed higher burden-and despair-related content and a broader range of high-arousal and affiliative emotions, including anxiety and happiness in particular sections. These findings are broadly compatible with prior work on gender differences in emotional expression, interpersonal framing and suicide-note content, although previous findings regarding gender and burdensomeness have not always been uniform ^14,20–22^. Importantly, these differences were expressed not only through overall theme prevalence but also through narrative placement: female-authored notes more strongly concentrated love and gratitude in concluding sections, whereas male-authored notes showed a more distributed presence of apology and shame. Explanations involving social obligation, self-reproach, interpersonal repair, gendered emotional expression or affiliative closure are plausible, but not directly tested here. The more defensible conclusion is that gender is associated with differences in the organization of final written communication, visible only when notes are modeled sequentially rather than as static texts.

The present findings have implications for suicide-note research and for the broader use of naturalistic text in behavioral science. Suicide notes are valuable because they are rare records composed close to death, but they are not transparent samples from all suicide decedents. They are produced by a selective subgroup and organized through communicative structures that shape what is expressed, where it appears and how it unfolds. This perspective is consistent with sociological accounts of suicide as embedded in social relationships and collective norms ^1^, as well as interpersonal theories emphasizing burdensomeness, belongingness and social disconnection as central to suicidal desire ^11,13^. The present study adds a distinct empirical point: relational, moral, emotional and practical dimensions are not only present in suicide notes; they are organized across the internal progression of the message.

Methodologically, the section-based framework illustrates how modeling naturalistic text as sequential communication can reveal behavioral patterning that whole-document summaries obscure. Although developed here for suicide notes, the same principle may extend to other consequential communications produced under emotionally constrained conditions, including farewell messages, crisis communications and digital final messages. Future research should test whether comparable organizational features appear in prospective communication streams before suicidal behavior. The present data are retrospective and postmortem, so they do not justify clinical prediction from note structure alone. Nevertheless, they suggest that models of suicide-related communication may benefit from considering organization as well as content.

Several limitations qualify these conclusions. First, the study is observational and registry-based, so the reported associations should not be interpreted causally. The analyses identify correlates of note leaving and structural regularities within notes, but cannot determine why individuals wrote notes, whether particular structures reflect specific intentions or how these communications relate to unobserved psychological states. Second, cultural, cohort and institutional factors may shape both note leaving and linguistic expression, limiting direct comparability with findings from other countries or periods. This issue may be especially relevant in South Korea, where suicide occurs within a sociocultural context marked by distinctive pressures and norms around family responsibility, obligation, social harmony and shame ^23,24^. Third, although the registry is unusually large and nationally comprehensive, several contextual variables rely on administrative coding, and information about the timing, recipient and interpersonal interpretation of pre-death communication was limited.

Additional methodological limitations also warrant consideration. Despite procedures used to distinguish raw notes from investigator-mediated summaries, residual heterogeneity in note recording, transcription, translation, paraphrasing or documentation cannot be fully excluded. The three-section framework is analytically useful but remains a heuristic segmentation strategy rather than a direct measure of psychologically validated narrative boundaries. Similarly, thematic and sentiment representations depend on the dictionaries, classifiers and thresholds used to define linguistic features. Future work should test alternative segmentation strategies, human annotation, contextual language models and cross-cultural validation while preserving interpretability.

Suicide notes are often treated as direct windows into suicidal experience. The present findings suggest a more qualified and scientifically useful view: they are selective and internally organized behavioral traces, produced by a non-random subset of suicide decedents and structured through recurrent communicative patterns. They contain distress, but they also organize apology, affiliation, responsibility and practical closure across the progression of a final message. The study therefore contributes to suicide research and to a broader behavioral-science question of how naturalistic texts should be used as evidence when both representativeness and internal organization are at stake.

## METHODS

### Ethics Statement

This study involved secondary analysis of anonymized registry records and suicide notes provided by the Korea Foundation for Suicide Prevention. It did not involve living human participants, and no personally identifying information was accessible to the research team. The study protocol was reviewed by the Institutional Review Board of the Korea Advanced Institute of Science and Technology (KAIST), which determined that the study was exempt from full ethical review. (Exemption Confirmation No. IRB-23-154).

### Demographic Data

The dataset used in this study consisted of nationwide police investigation reports of suicide cases collected and organized by the Korean Foundation for Suicide Prevention (KFSP). The data was collected between 2013 and 2020 for the specific purpose of empirically studying suicide cases in Korea. A total of 102,922 cases were compiled nationwide from police stations across Korea and the data consists of a wide range of demographic variables, suicide-related variables, health-related variables, variables related to life stressors, and suicide notes.

The dataset included demographic variables (age group, gender, marital status, cohabitation status, job type, employment type), suicide-related characteristics (murder suicide, group suicide, prior attempts, method, location, discovery context, warning signs, and putative cause), life stressors (economic, job-related, family, and relationship problems), health indicators (physical and mental health conditions, psychiatric history, alcohol consumption at death), and additional qualitative fields describing psychological symptoms and specific stressor details (Proportions plots are seen in Extended Data 7-10).

### Suicide Notes

Among the 102,922 cases, 38,044 cases contained suicide notes. Personally-identifying information (e.g., names, addresses, national identification numbers) had been anonymized by data custodians beforehand. For 582 cases, note content was not disclosed in the dataset or contained multiple languages; thus 37,462 notes were eligible for text processing. When multiple notes were recorded for a single death, they were concatenated and treated as one document.

### Identification of Raw versus Summarized Suicide Notes

The registry contained both verbatim suicide notes and investigator-generated summaries or paraphrased excerpts. Because note type was not explicitly labeled, and summarized notes may differ linguistically from original notes, we developed a weak-supervision pipeline to probabilistically classify notes as *raw* (verbatim content written by the deceased) or *summarized* (investigator-mediated paraphrase or excerpt).

### Rule-based weak labeling

We implemented 27 labeling functions to detect linguistic and structural features characteristic of investigator summaries in police documentation. These functions captured reportative constructions (e.g., equivalents of “it was reported that…”), meta-report expressions (e.g., “a suicide note was found”), third-person narrative shifts, structured header-style formats (e.g., itemized summaries), message-transcript indicators (e.g., references to text or messaging content followed by reporting verbs), date-leading summary formats (e.g., timestamps followed by paraphrase content), and written-summary constructions (e.g., equivalents of “it was written that…”). Each function emitted a vote (raw = 0, summarized = 1) or abstained. For notes receiving at least one non-abstaining vote, a weak label was assigned by majority vote. A rule-confidence score was defined as the absolute vote imbalance divided by the total number of votes for that note, and only notes with rule-confidence ≥ 0.34 were retained as weakly labeled training examples. Notes without confident weak labels were excluded

### Feature representation and classifier training

We trained a supervised classifier on the weakly labeled subset using character-level n-grams (3–5 characters) weighted by term frequency–inverse document frequency (TF–IDF) to capture stylistic differences beyond explicit rule patterns. A logistic regression classifier with L2 regularization (scikit-learn) was fit with class-weight balancing, and probability calibration was performed using cross-validated Platt scaling (CalibratedClassifierCV). Raw note length was not included as a feature to reduce discrimination based solely on document length. Model predictions were compared against weak labels in a held-out subset as an internal consistency check; because weak labels may contain noise, these checks were interpreted as indicators of model stability rather than gold-standard performance.

### Probability blending and final classification

For each note, the final probability of being summarized combined the calibrated classifier probability (P_model) with a rule-derived pseudo-probability (P_rule) based on rule-confidence. The blended probability was computed as 0.7·P_model + 0.3·P_rule. For notes without investigator-style context cues—where rule-based labeling functions typically abstained and yielded pseudo-probabilities near 0.5—the blending weights were adjusted to 0.35·P_model + 0.65·P_rule. This weighting scheme effectively shrinks predictions toward 0.5 in cue-absent cases, reducing overconfident model-based classifications in linguistically ambiguous notes. Using a classification threshold of 0.50, 25,992 notes were classified as raw and 11,470 as summarized. All primary linguistic analyses were conducted on the subset classified as raw.

### Manual validation and internal robustness analyses

To assess criterion validity, we manually labeled an independent set of 1500 notes as raw or summarized and compared these labels with model predictions. Agreement between manual labels and model

We additionally conducted several internal robustness analyses. Across classification thresholds ranging from 0.45 to 0.60, the proportion of notes classified as raw varied from 69.01% to 70.05%, indicating limited sensitivity of the overall class distribution to moderate changes in the decision threshold. Under 200 full-size bootstrap resamples with the fitted classifier held fixed, the mean proportion classified as raw was 69.42%, with a standard deviation of 0.25 percentage points and a percentile-based 95% interval of 68.97% to 69.85%.

In counterfactual cue-removal analyses, removing explicit investigator-style summary cues reduced the summary-classification score among notes classified as summarized by an average of 0.259. Conversely, removing predefined raw-associated cues increased the summary-classification score among notes classified as raw by an average of 0.048. These results indicated that the classifier responded more strongly to investigator-style documentation markers than to the smaller set of raw-associated lexical cues.

The predicted score distributions showed very low mixture overlap, with an estimated overlap of approximately 0.000005. Final classifications agreed with non-abstaining weak labels for 99.62% of 24,230 notes, and the Brier score relative to these weak labels was 0.0035. Across five bootstrap-refitted models, mean agreement with the majority classification was 99.55%, and 98.44% of notes received unanimous classifications across all models. Because the weak labels were used to construct the classifier’s training data, agreement and probability error relative to these labels were interpreted as measures of internal consistency rather than independent classification validity. Together, the manual validation and robustness analyses indicated that the classification procedure showed substantial agreement with human annotation and high stability across thresholds, bootstrap resampling, cue-removal tests and model refitting.

### Within-note segmentation

Each raw note was segmented into three consecutive (Introduction, Body, Conclusion). When the number of sentences was not divisible by three, remainder sentences were appended to the Conclusion segment. To avoid empty sections, section-based analyses were restricted to raw notes with ≥3 sentences (n = 18,635). Sentence splitting used rule-based punctuation heuristics implemented in the kss Python module.

Because any fixed partition of free text is necessarily heuristic, we assessed whether the main positional findings depended on this specific segmentation choice. In addition to the primary three-section analyses, we repeated the analyses using four-section partitions, restricting to longer notes with at least 5 sentences, and sentence-level models based on each sentence’s relative position within the note. The sentence-level analyses showed the same broad positional trends as the primary section-based results, and the alternative section-based specifications yielded qualitatively similar patterns. Together, these checks indicate that the observed placement and sequencing effects reflect robust within-note organization rather than an artifact of a particular partitioning rule (See Extended Fig. 3-6).

### Demographic Analyses

Logistic regression was used to identify factors distinguishing suicide note-leavers from non-leavers. All predictors were categorical, with reference categories defined as the most prevalent level within each variable. Cases with missing responses were excluded; however, the extent of missingness varied widely across predictors, necessitating a systematic approach to variable inclusion. To balance model complexity with sample retention, we applied a data-driven procedure that quantified the trade-off between adding predictors and the resulting number of complete cases.

This analysis revealed inflection points at which additional variables sharply reduced the analyzable sample, defining 8 progressively inclusive subsets of predictors. Logistic regression models were fit for each subset, and evaluated using likelihood ratio chi-squared statistics (model fit), Nagelkerke’s pseudo-R2 (explained variance), classification accuracy, sensitivity, specificity, and effective sample size (*see Extended Table 1*).

Excluding overly restrictive models that severely reduced the analytic sample, Subset 3 was selected as the optimal compromise between explanatory power and classification performance. While more inclusive models yielded marginal improvements in accuracy and variance explained, these came at the expense of drastically reduced sample sizes, limiting generalizability. By contrast, Subset 3 retained a large sample (*N≈70,000*), explained a comparatively substantial proportion of variance (*Nagelkerke’s R2≈12.3%*), and achieved the highest sensitivity (*≈38%*). All subsequent interpretations are based on the results from this model. Regression analyses were conducted in R (version 4.4.1).

### Construction of theme matrices

The thematic structure of each suicide note was quantified using a predefined set of five thematic categories based on previous suicide note studies ^5,7^: Sorry and Shame, Love and Gratitude, Burden, Despair, and Post-mortem Affairs. After each note was divided into three consecutive textual sections, the keywords in each section were extracted using a cosine similarity-based approach with KeyBERT ^27^. These words were then scored for thematic presence using a theme-relevant word-dictionary approach (*see Extended Table 2*). This resulted in a 15-dimensional feature vector for each note (5 themes × 3 sections), representing the relative and explicit presence of each theme across the note’s narrative progression.

### Construction of sentiment matrices

To explore the sentiments associated in the text, a pre-trained KcELECTRA Korean language model ^28^ was used to extract a subset of 15 emotions that were associated with suicidality based on psychosocial theories of suicide ^12,13,26,29,30^. The classifier extracted sentiments in each section of suicide notes after the notes were split into the 3 given sections. Confidence scores (0-1) were assigned to the sentiments across each sentence in a multi-label format, with a threshold of 0.7 applied. For every section we quantified the prevalence of 15 sentiment categories (*Despair, Defeat, Exhaustion, Anxious, Disappointment, Anger, Hatred, Resentment, Sadness, Sorrow, Gratitude, Affection, Happiness, Relief, and Neutral*), producing a 45-dimensional vector per note (3 sections × 15 sentiments).

### Clustering thematic and sentiment trajectories

Clustering was first performed on a PCA-reduced version of these matrices using the K-Means algorithm ^31,32^. A threshold for the minimum explained variance was set to 50% and the goodness of separation from the K-means algorithm was measured using silhouette scores. Based on these constraints, a range of 0-20 PCA clusters and K-means clusters were tested accordingly to determine the optimal separability (Silhouette score) while maintaining a threshold explained variance. As a result, the theme matrices were reduced to 5 dimensions and K-means clusters were set to 15 based on the silhouette score and thematic interpretability considerations. This yielded in an explained variance of 0.53 and a silhouette score of 0.49. For the sentiment matrices, the vectorized notes were reduced to 6 dimensions and K-means clusters were set to 10 based on the silhouette score and thematic interpretability considerations. This yielded in an explained variance of 0.51 and a silhouette score of 0.22. The algorithm was initialized with 10 random seeds to mitigate local minima and the final solution was selected based on minimal within-cluster sum of squares. These clusters were subsequently labelled based on the dominant cluster centroids across the clusters (see Extended Table 3). Because centroids are reconstructed from PCA space, occasional values slightly outside 0,1 occur; we therefore interpret them relationally (higher vs. lower; rising vs. falling) rather than as absolute probabilities.

### Sequential transition analysis of within-note progression

For each theme or sentiment present in a given section, we estimated the probability that another theme or sentiment appeared in the subsequent section. Specifically, for every pair of states *i* (in Section t) and *j* (in Section t+1), we computed the proportion of notes in which state *j* appeared in the next section among notes in which state *i* appeared in the current section. This yielded a square transition matrix summarizing conditional co-occurrence patterns from Section 1 to Section 2 and from Section 2 to Section 3, separately for themes and sentiments.

Because some themes and sentiments are more prevalent overall than others, raw transition probabilities may be inflated by base-rate effects. To account for this, we normalized transition probabilities using a lift-based measure, commonly used in association-rule mining to quantify whether two states co-occur more often than expected from their marginal frequencies ^33^. For each transition, the conditional probability of observing a given state in the next section was divided by the marginal prevalence of that state in that section across all notes. Lift values greater than 1 indicate that a state appears more frequently following a given prior state than would be expected based on its overall prevalence, whereas values below 1 indicate relative underrepresentation.

For each comparison, we computed the difference in lift (ΔLift) between the focal group and the comparison group. Positive ΔLift values indicate transitions that are more strongly enriched in the focal group, whereas negative values indicate transitions that are relatively more characteristic of the comparison group. ΔLift matrices were computed separately for transitions from Section 1 to Section 2 and from Section 2 to Section 3, and separately for themes and sentiments. Statistical significance of ΔLift values was assessed using nonparametric permutation testing. Group labels (e.g., gender or age group) were randomly reassigned across notes while preserving original group sizes, and ΔLift matrices were recomputed for each permutation. This procedure generated a null distribution of ΔLift values for every transition cell.

Two-sided p-values were calculated as the proportion of permutations in which the absolute permuted ΔLift value equaled or exceeded the observed value. To address the large number of transitions tested simultaneously, we additionally implemented an optional max-statistic correction, in which the maximum absolute ΔLift value across all transition cells was recorded for each permutation. This approach provides family-wise error rate control while preserving the dependence structure among transitions. Unless otherwise stated, results are reported using cellwise permutation p-values, with max-statistic correction used as a robustness check (All ΔLift values are seen in Supplementary Table 4-15).

### Logistic regression analysis

To estimate how section-specific themes and sentiments were associated with age and recorded sex, we fitted separate multivariable binary logistic regression models for the thematic and sentiment features. Age was categorized as ≤18, 19–34, 35–49, 50–64 and ≥65 years, and recorded sex was coded as female or male. For each age category, we created a binary outcome comparing the target age group with all other age groups and included recorded sex as a covariate. For the recorded-sex analysis, we modelled male versus female and included age group as a categorical covariate. All section-specific features within the corresponding thematic or sentiment domain were entered simultaneously. Coefficients are reported as log odds ratios with 95% confidence intervals. To account for multiple hypothesis testing, two-sided p-values were adjusted using the Benjamini–Hochberg false-discovery-rate procedure within four prespecified families: theme-by-age tests (75 comparisons), theme-by-recorded-sex tests (15 comparisons), sentiment-by-age tests (225 comparisons) and sentiment-by-recorded-sex tests (45 comparisons). The five one-versus-rest age models were pooled within the corresponding age correction family. Statistical inference and significance labels were based on the FDR-adjusted p-values, with *FDR-adjusted p < 0.05, **FDR-adjusted p < 0.01 and ***FDR-adjusted p < 0.001.

## Data availability statement

The data used in this study were provided by the Korea Foundation for Suicide Prevention (KFSP) under a data use agreement. Because the dataset contains sensitive information related to suicide deaths, it cannot be made publicly available.

## Code availability statement

The custom code used for text preprocessing, thematic and sentiment classification, section-based segmentation, clustering, and statistical modeling is available from the corresponding author upon reasonable request.

## ACKNOWLEDGEMENTS

This research was supported and funded by the Korea Foundation for Suicide Prevention under the designated grant code (KFSP23-02).

## AUTHOR CONTRIBUTIONS

J.B.P., A.A, S.Y.O., and J.J. conceived and designed the study. J.B.P., A.A. and performed data processing, computational analysis and statistical modeling. Y.H.H., S.J.L. and S.K. provided the dataset, and contributed domain expertise and feedback on suicide research, data interpretation, and the analyses. J.B.P., A.A, J.J. wrote the manuscript with input from all authors. J.J. supervised the project. All authors reviewed and approved the final manuscript.

## Extended Data

**Extended Data Fig. 1.**
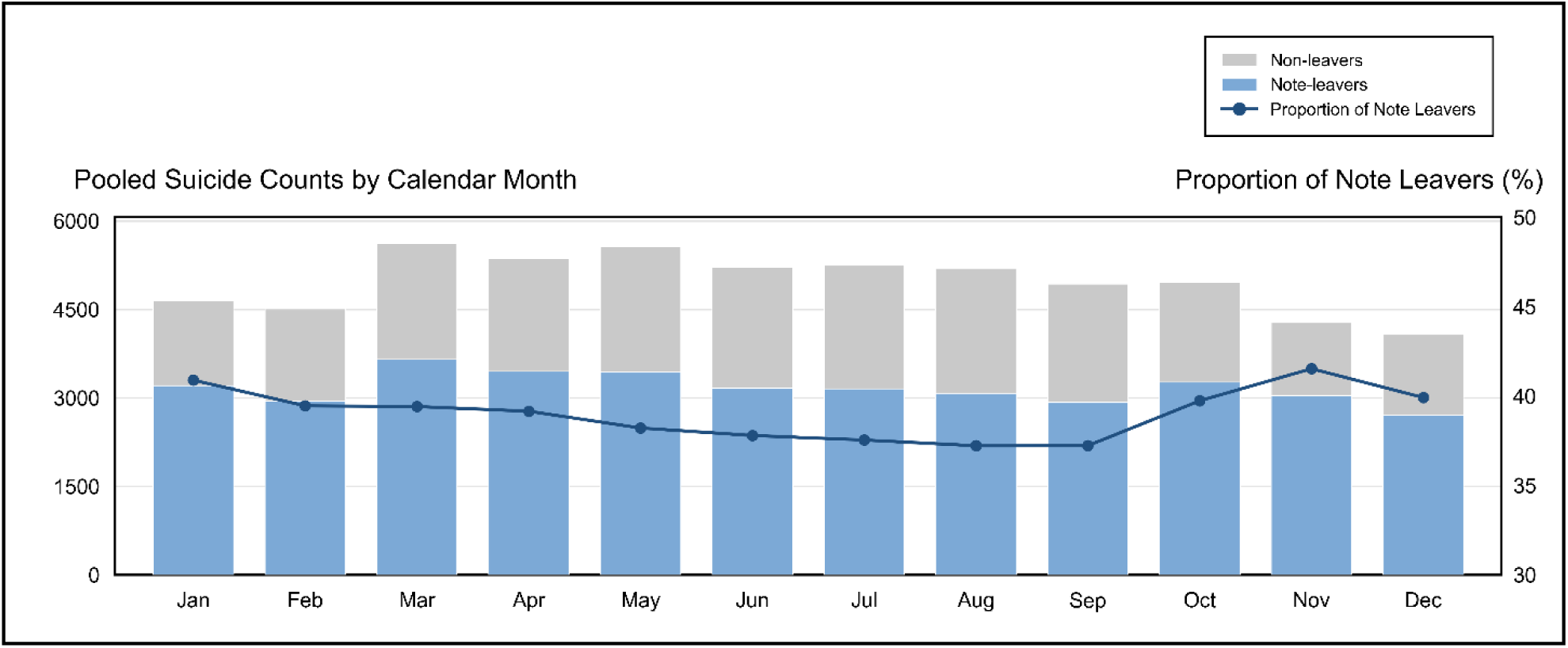
Monthly suicide counts stratified by suicide note status (2013–2020). Monthly suicide deaths aggregated across the study period are shown separately for individuals who left suicide notes (blue) and those who did not (gray). Both groups exhibit similar seasonal dynamics, with peaks in early months of the calendar year. The proportion of suicide deaths accompanied by notes varies across months and tends to increase toward the end of the year.

**Extended Data Fig. 2.**
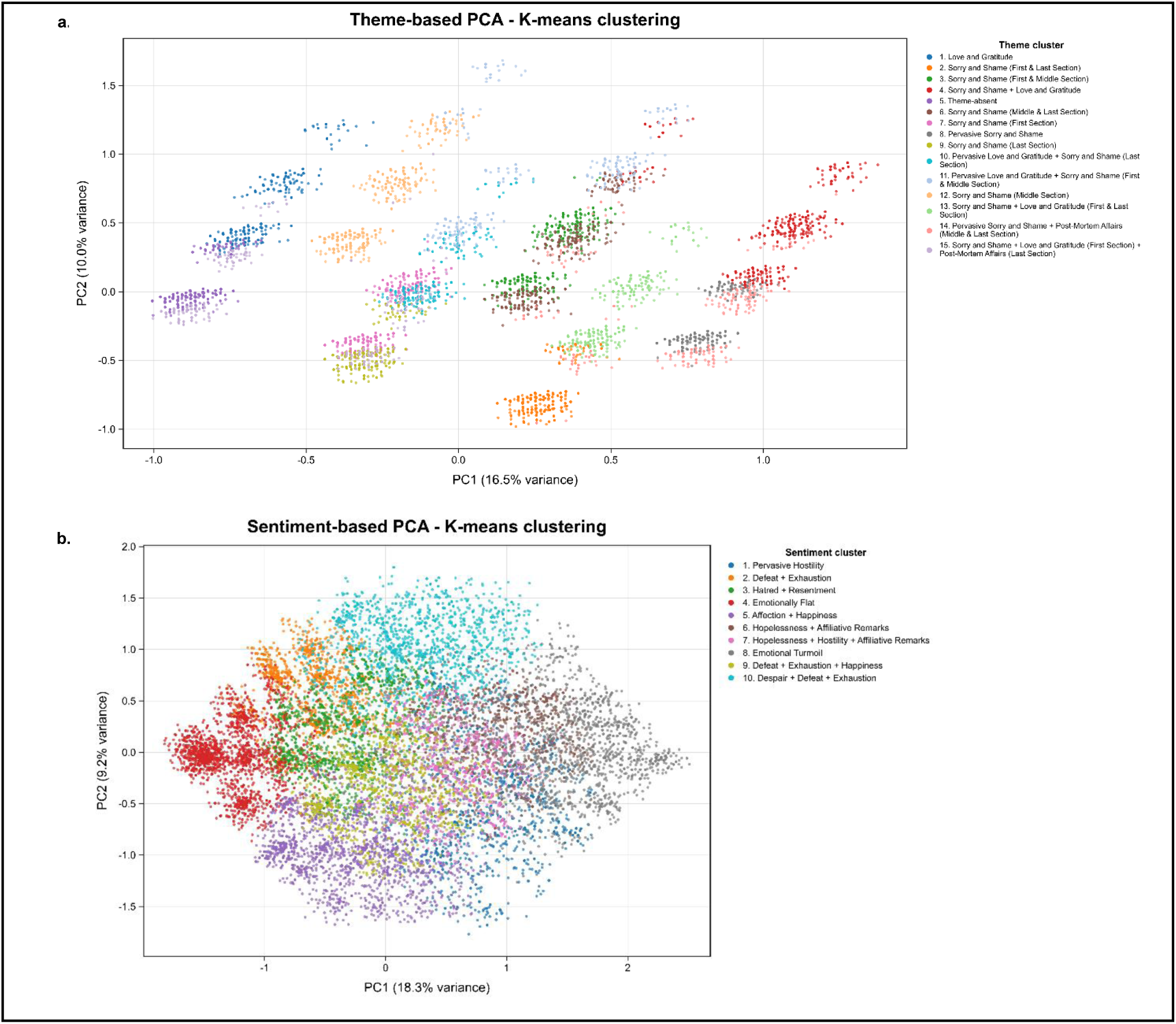
PCA projections of thematic and sentiment clusters. a. PCA projection of suicide notes based on theme matrices derived from section-based representations. Each point represents a suicide note projected into two principal component dimensions. Fifteen thematic clusters were identified using K-means clustering based on silhouette score and interpretability criteria.b. PCA projection of suicide notes based on sentiment matrices extracted from sectioned notes. Ten sentiment clusters were identified. Cluster descriptions are provided in Supplementary Data Table 3.

**Extended Data Figure 3.**
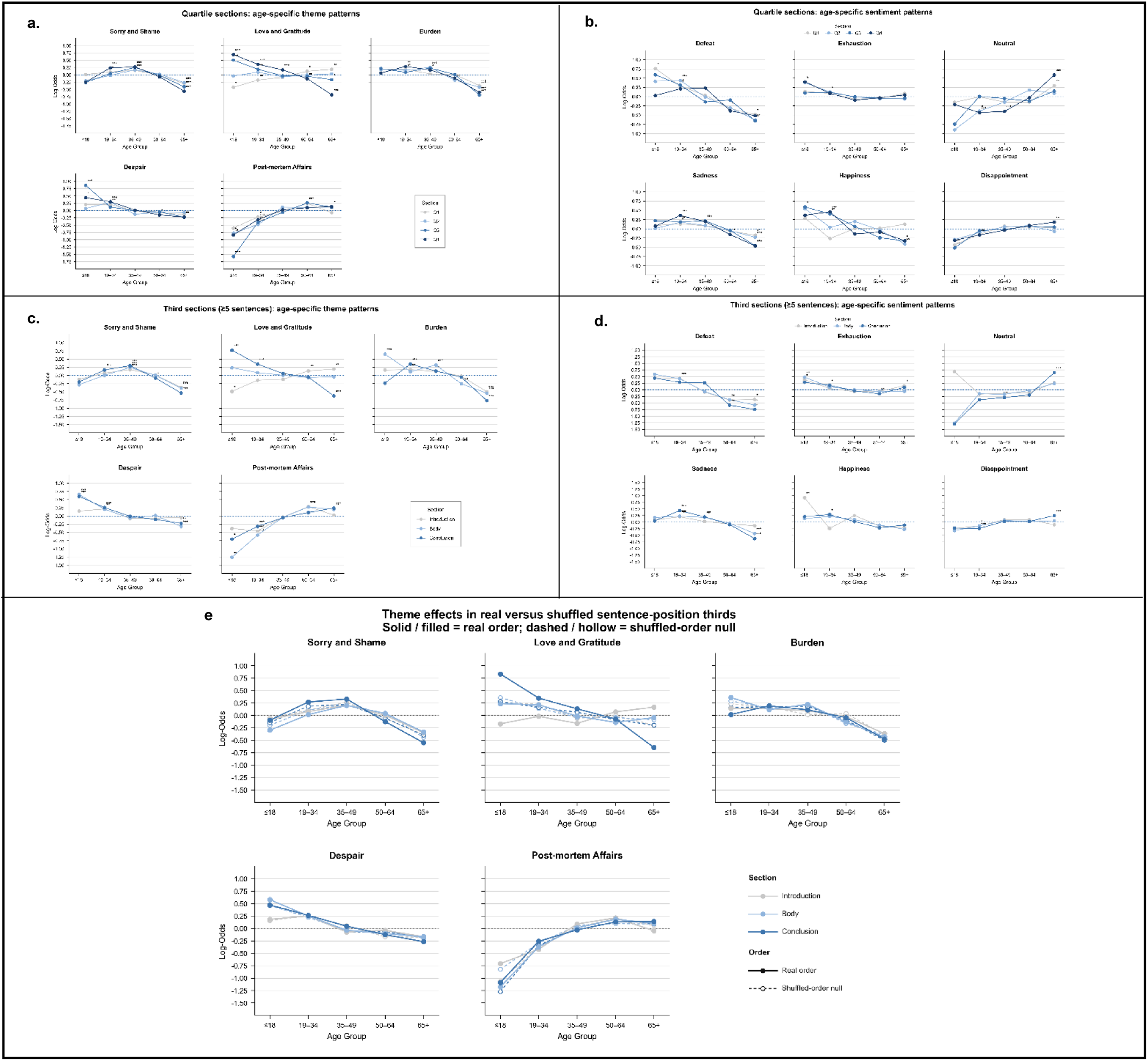
Section-Robustness Check with Quartile and Longer Notes for Age Groups (More than 5 sentences). –. **a.)** presents the log odds ratio (with CIs) of theme sections across age, **b.)** presents log odds ratios of sentiment sections across age when notes are split into quartiles. **c.)** presents the log odds ratios of theme sections and **d.)** presents log odds ratio of sentiment sections when notes that are longer than 5 sentences are used for analysis. **e.)** presents the logs odds ratios of themes across age groups when notes are compared in their real order (solid lines) and shuffled order (dashed lines), providing validation for significant section-based effects seen with love and gratitude in the results. The log odds ratio are presented across sections and significances are labelled based on p<0.05, 0.01, 0.001. Results show comparable results to the original section-based results.

**Extended Data Figure 4.**
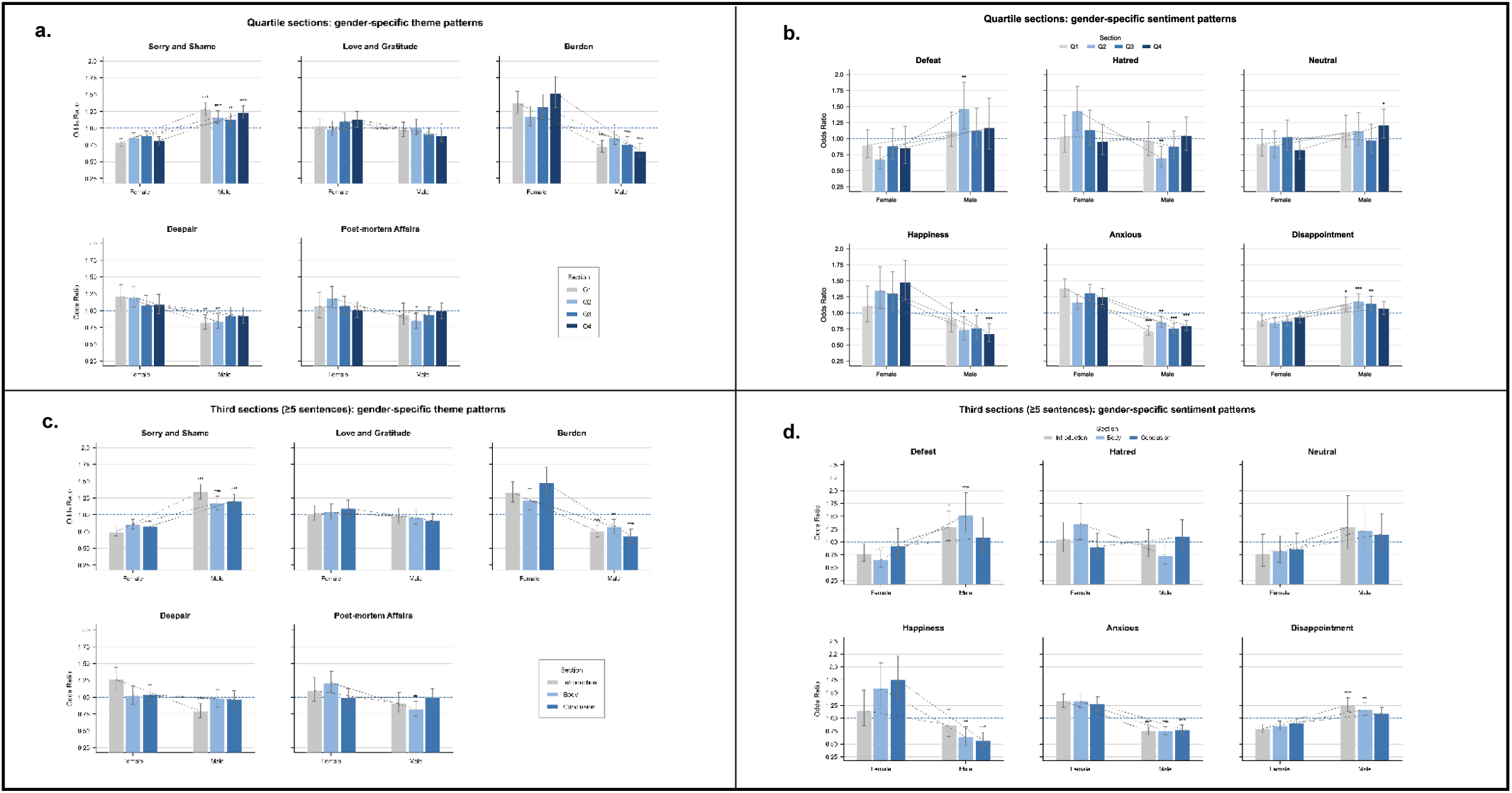
Section-Robustness Check with Quartile and Longer Notes for Gender (More than 5 sentences). –. **a.)** presents the log odds ratio (with CIs) of theme sections across gender, **b.)** presents log odds ratios of sentiment sections across gender when notes are split into quartiles. **c.)** presents the log odds ratios of theme sections and **d.)** presents log odds ratio of sentiment sections when notes that are longer than 5 sentences are used for analysis. The log odds ratio are presented across sections and significances are labelled based on p<0.05, 0.01, 0.001. Results show comparable results to the original section-based results.

**Extended Data Figure 5.**
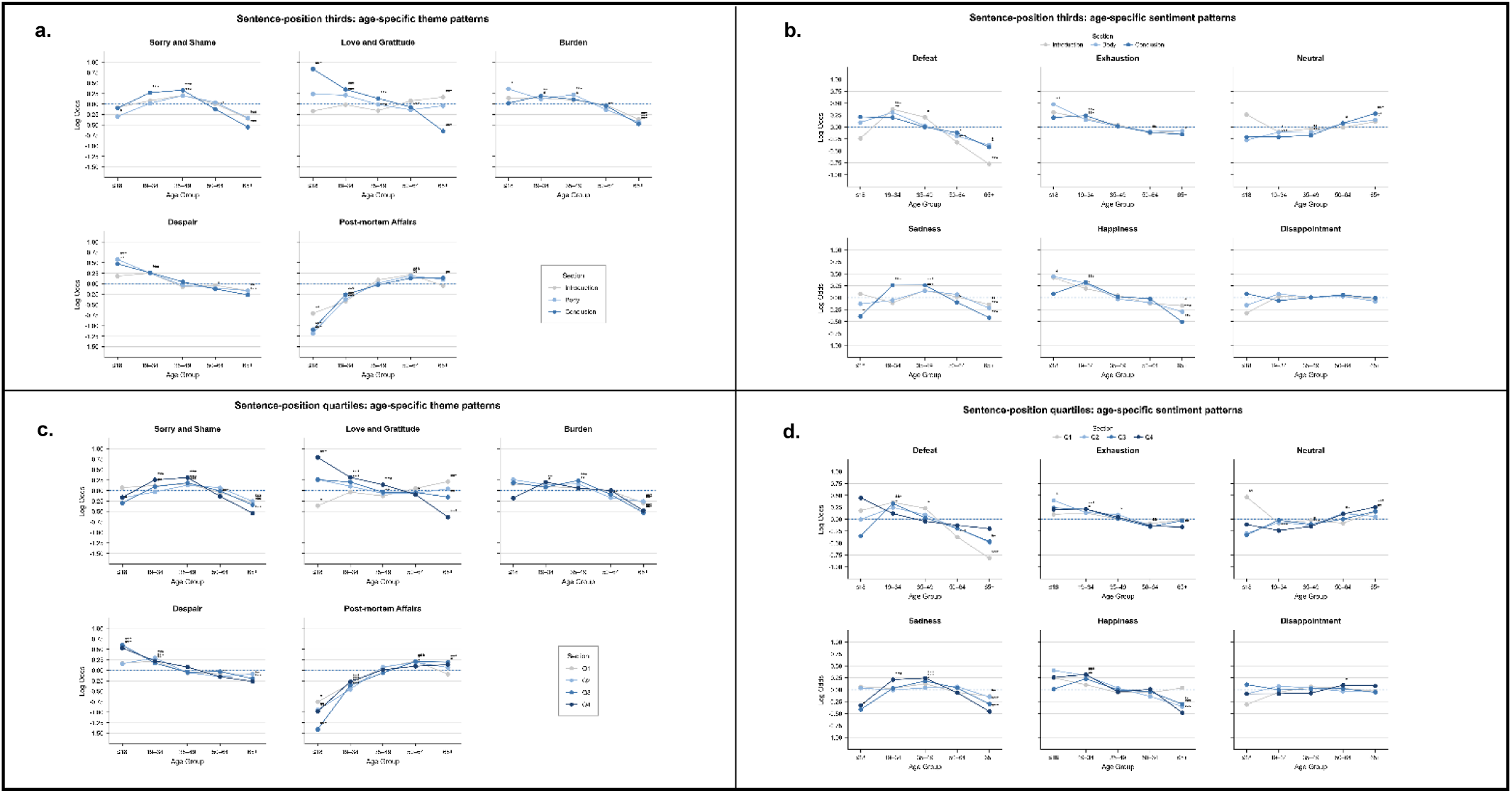
Sentence-position robustness analyses of age-related thematic and sentiment structure in suicide notes for age groups. a, b, Sentence-level theme (a) and sentiment (b) analyses using a three-section division of each note into opening, middle and ending segments. c, d, Parallel analyses using a four-quartile division of each note into Q1–Q4. For each note, sentence-level theme and sentiment flags were aggregated within each section, indicating whether a given feature appeared in at least one sentence in that segment. Points represent log odds ratios from one-versus-rest logistic regression models predicting membership in each age group while adjusting for sex; lines connect estimates for the same section across age groups. Values above zero indicate greater relative occurrence in the focal age group, and values below zero indicate lower relative occurrence. The three-section and quartile analyses show broadly convergent age-related structuring of final communications, supporting the robustness of the section-based findings to alternative sentence-position definitions. Significance markers indicate two-sided Wald tests: P < 0.05, P < 0.01, P < 0.001.

**Extended Data Figure 6.**
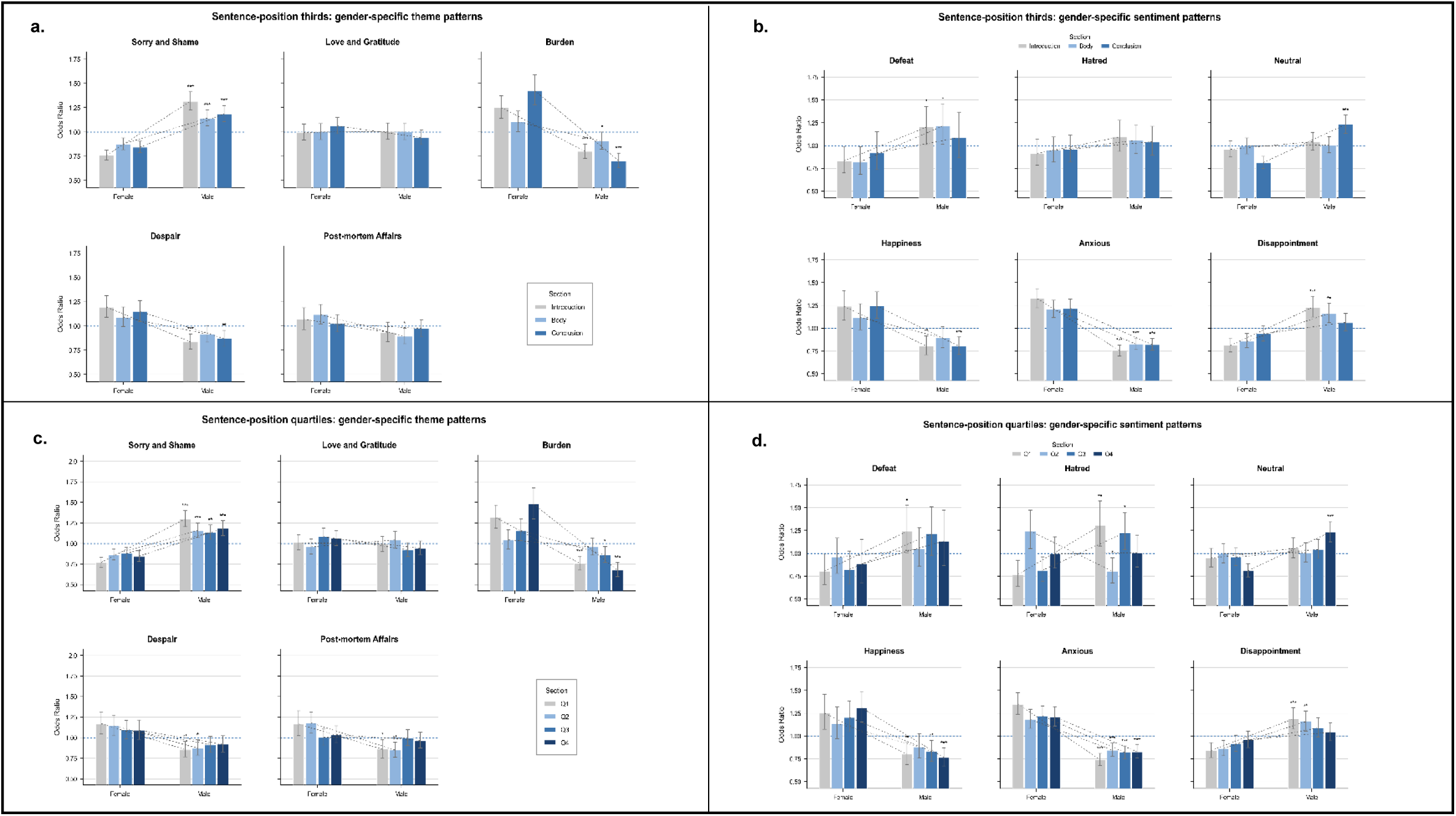
Sentence-position robustness analyses of age-related thematic and sentiment structure in suicide notes for gender. a, b, Sentence-level theme (a) and sentiment (b) analyses using a three-section division of each note into opening, middle and ending segments. c, d, Parallel analyses using a four-quartile division of each note into Q1–Q4. For each note, sentence-level theme and sentiment flags were aggregated within each section, indicating whether a given feature appeared in at least one sentence in that segment. Points represent log odds ratios from one-versus-rest logistic regression models predicting membership in each age group while adjusting for sex; lines connect estimates for the same section across age groups. Values above zero indicate greater relative occurrence in the focal age group, and values below zero indicate lower relative occurrence. The three-section and quartile analyses show broadly convergent age-related structuring of final communications, supporting the robustness of the section-based findings to alternative sentence-position definitions. Significance markers indicate two-sided Wald tests: P < 0.05, P < 0.01, P < 0.001.

**Extended Data Figure 7.**
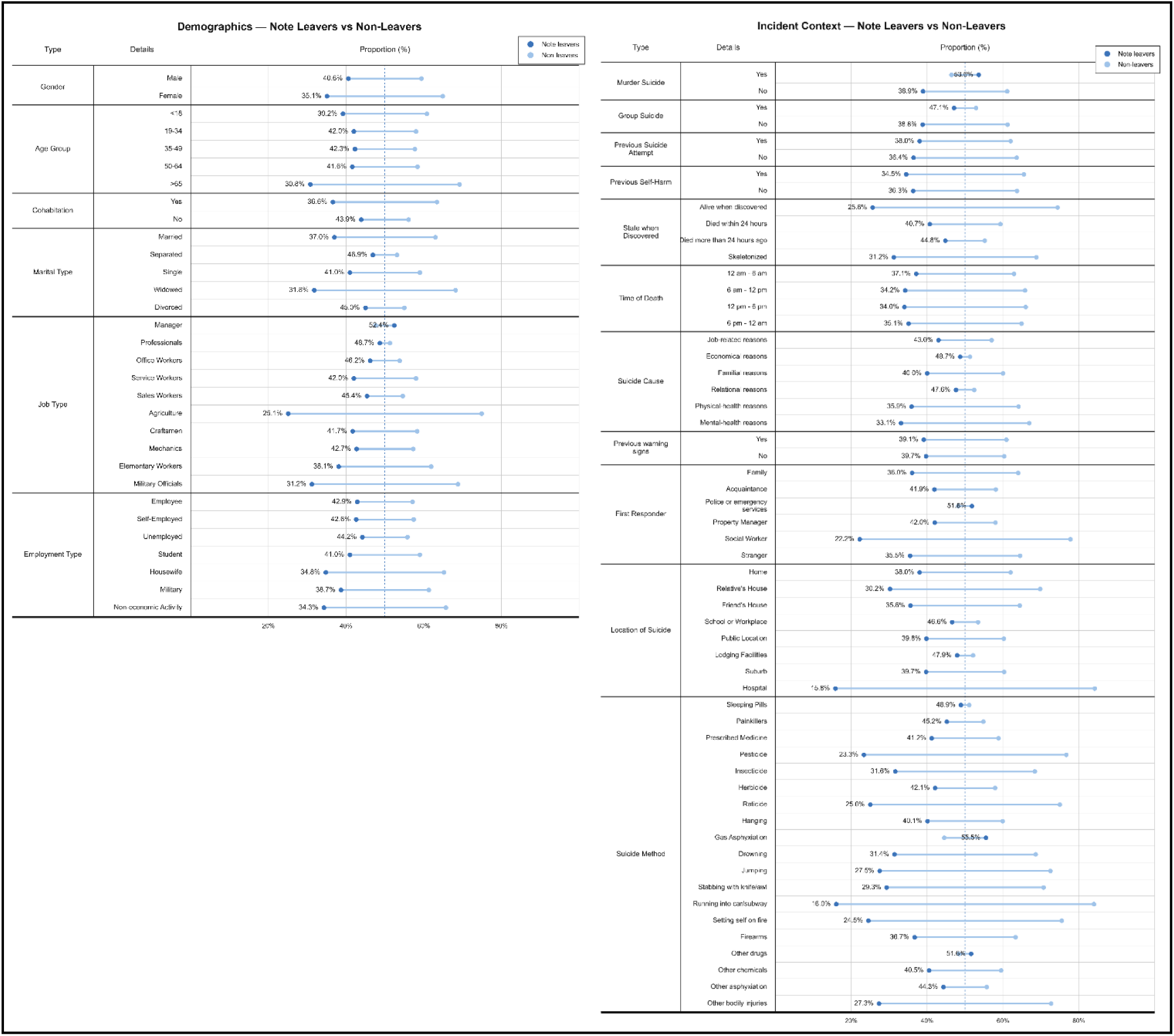
Sociodemographic characteristics and suicide context of note-leavers and non-leavers. Proportion of suicide cases accompanied by notes across demographic variables including gender, cohabitation status, age group, marital status, employment category and contextual variables including murder–suicide, group suicide, prior suicide attempts, prior self-harm, discovery state, time of death, suicide cause, first responder, location of suicide, and suicide method.

**Extended Data Figure 8.**
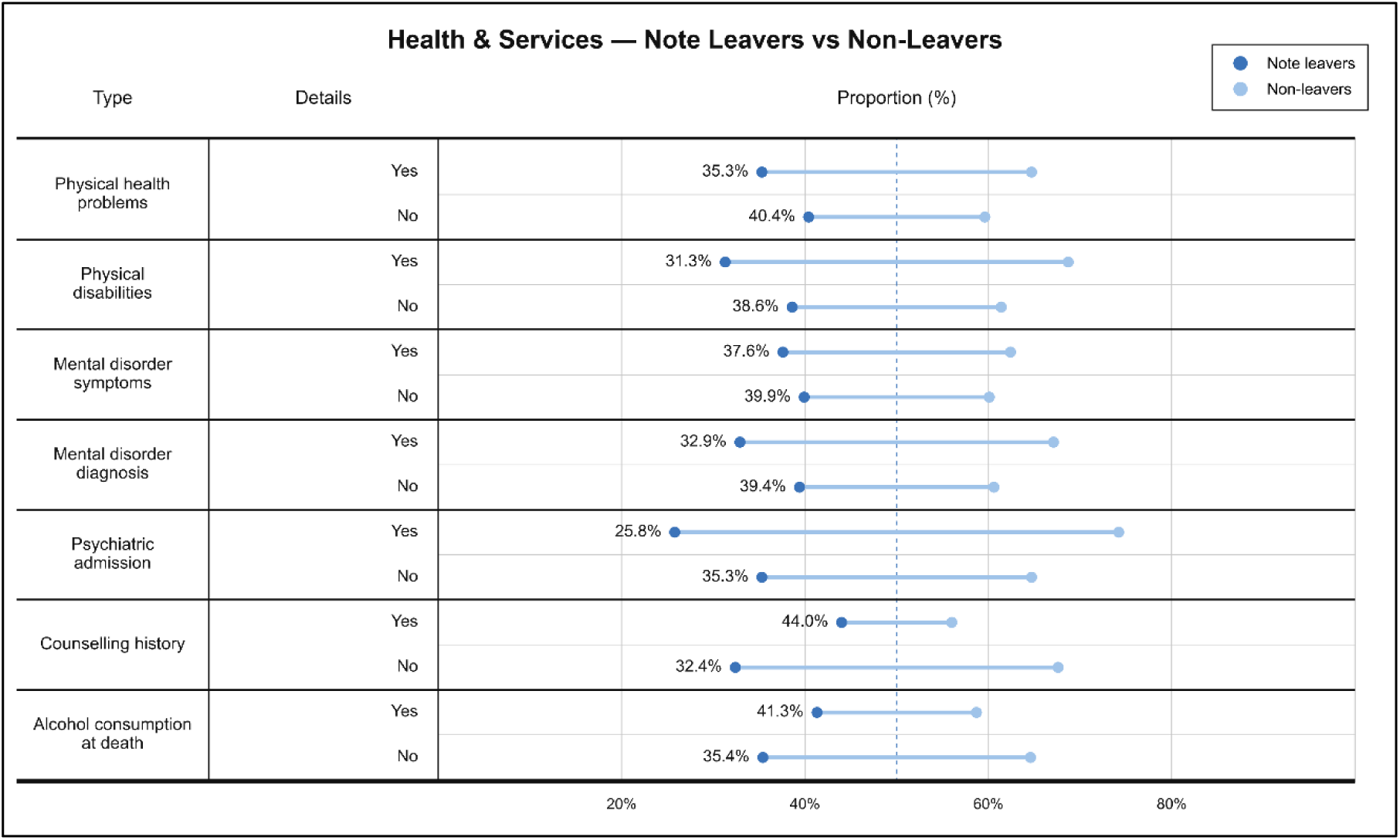
Physical health characteristics among note-leavers and non-leavers. Proportion of suicide note presence across health-related variables including physical health problems, physical disability, psychiatric symptoms, psychiatric diagnoses, history of psychiatric hospitalization, counseling history, and alcohol consumption at the time of death.

**Extended Data Figure 9.**
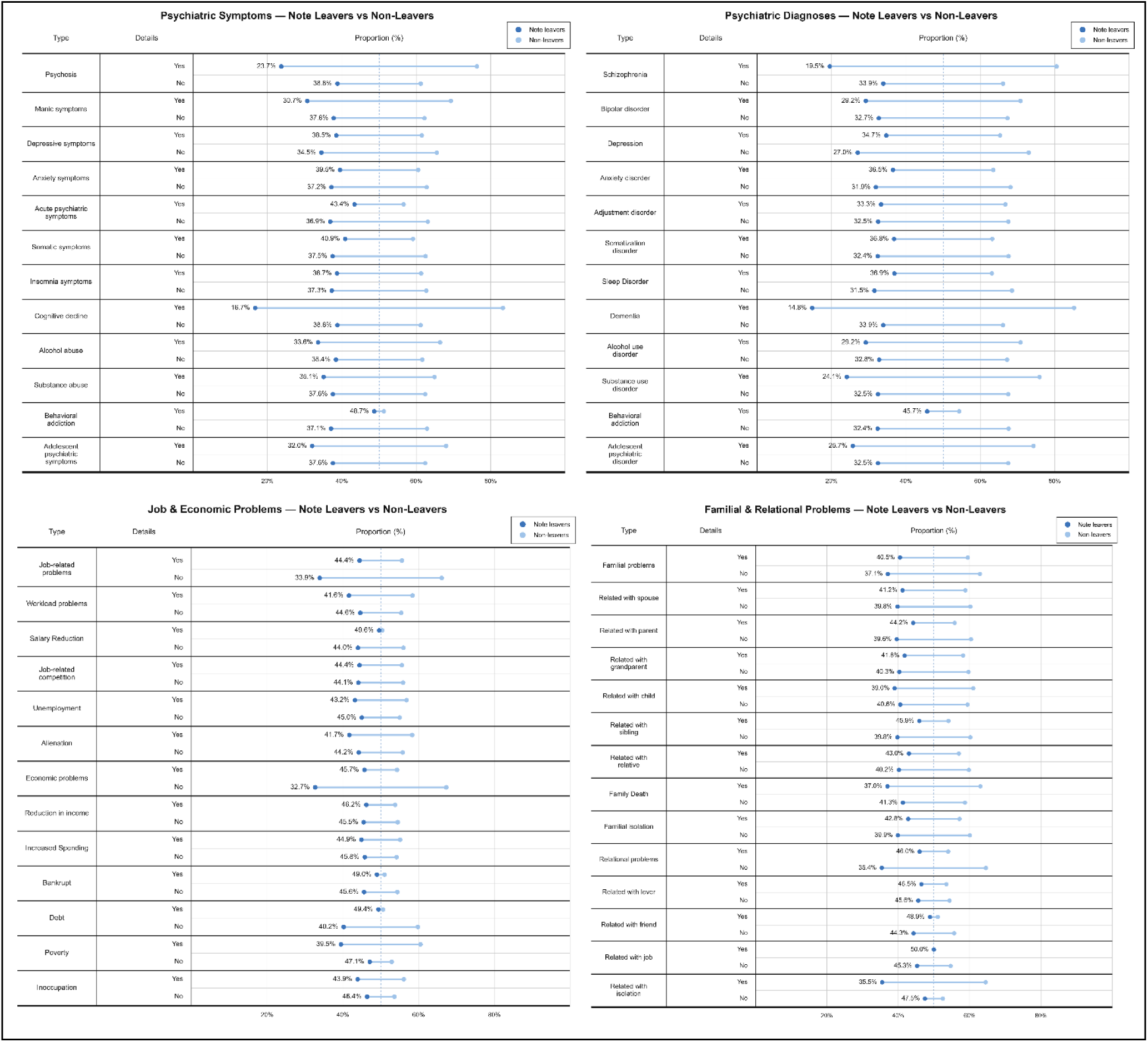
Detailed mental health characteristics and socioeconomic stressors among note-leavers and non-leavers. Proportion of suicide note presence across specific mental health-related and major socioeconomic stressors including job-related problems, financial difficulties, family conflict, and relational problems.

**Extended Data Figure 10.**
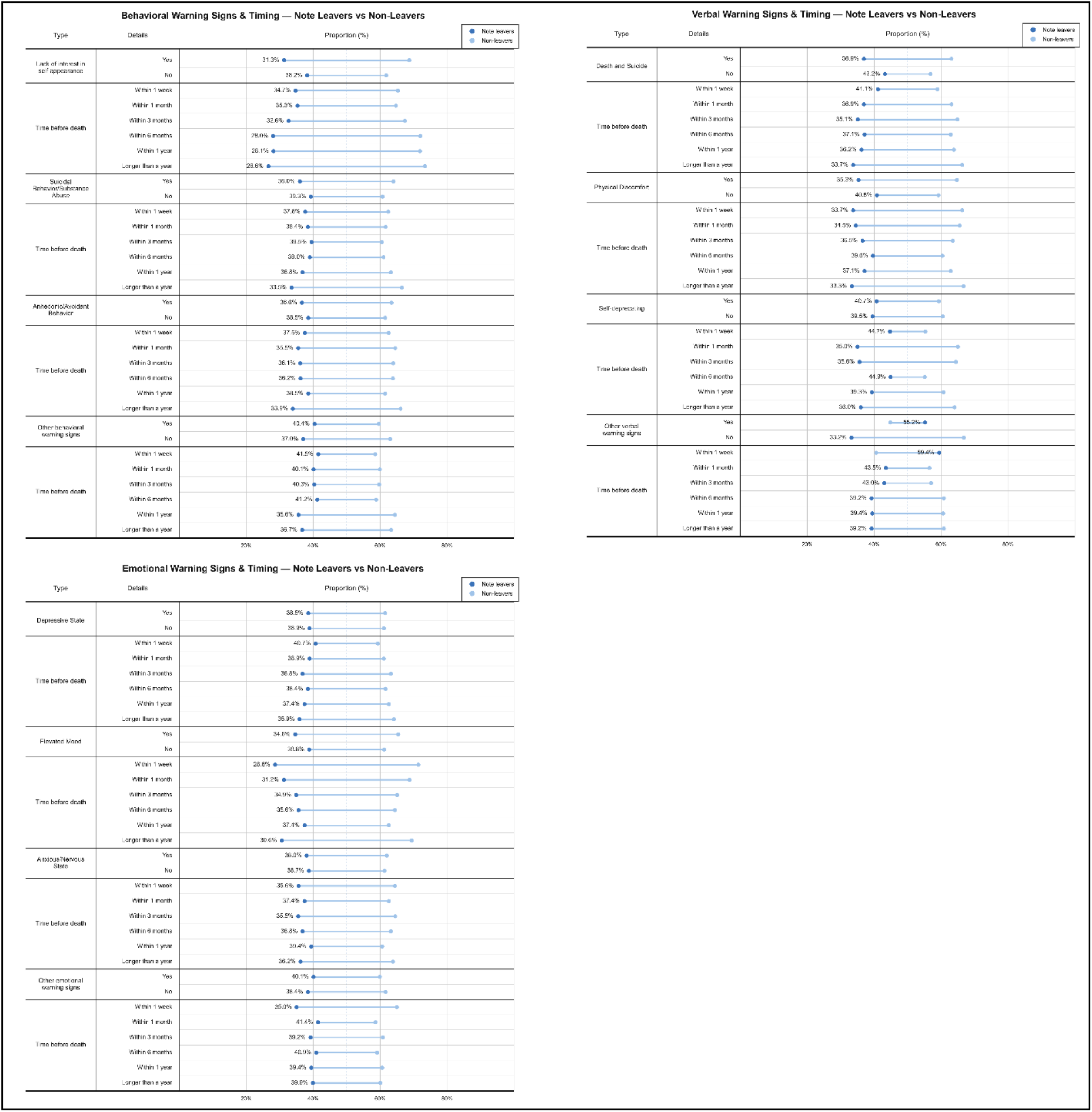
Prior warning signs among note-leavers and non-leavers. Distribution of previously reported warning signals prior to suicide, including verbal, behavioral, and emotional indicators. Bars represent the proportion of cases within each category.

