## Supplementary Data Table 1-15 for "Suicide notes as structured final communications: evidence from 100,000 deaths in South Korea"

**Supplementary Data Table 1:** Likelihood ratio chi-squared statistics (model fit), Nagelkerke's pseudo-R2 (explained variance), classification accuracy, sensitivity, specificity, and effective sample size of logistic regression models used in demographic comparisons.

| **Cuts** | **Chi square** | Degrees of Freedom | p_value | accuracy | sensitivity | specificity | nagelkerke_R2 | Total Count | actual_n0 | actual_n1 | predicted_n0 | predicted_n1 |
|---|---|---|---|---|---|---|---|---|---|---|---|---|
| Cut 1 | 5589.343341 | 61 | <0.00 | 63.03072 | 25.52646 | 86.99168 | 7.586222151 | 97086 | 59239 | 37847 | 79719 | 17367 |
| Cut 2 | 5510.070987 | 71 | <0.00 | 63.13621 | 25.26675 | 87.26174 | 7.766425145 | 93444 | 57080 | 36364 | 76985 | 16459 |
| Cut 3 | 6753.223381 | 92 | <0.00 | 64.7204 | 38.27369 | 82.4036 | 12.27605006 | 70922 | 42503 | 28419 | 52566 | 18356 |
| Cut 4 | 5000.055006 | 116 | <0.00 | 65.12983 | 31.40363 | 85.98829 | 11.42557324 | 56957 | 35192 | 21765 | 45191 | 11766 |
| Cut 5 | 4050.463323 | 150 | <0.00 | 65.21834 | 30.7088 | 86.33333 | 11.48378683 | 45938 | 28500 | 17438 | 36688 | 9250 |
| Cut 6 | 3143.507836 | 173 | <0.00 | 66.36972 | 29.83119 | 87.55722 | 12.62595296 | 32441 | 20534 | 11907 | 26334 | 6107 |
| Cut 7 | 1553.692234 | 192 | <0.00 | 67.04969 | 28.70712 | 88.51684 | 12.81711208 | 15839 | 10154 | 5685 | 13041 | 2798 |
| Cut 8 | 693.1913909 | 229 | <0.00 | 71.79433 | 31.83886 | 90.85555 | 18.91351382 | 4765 | 3226 | 1539 | 3980 | 785 |

**Supplementary Data Table 2:** Theme Matrix Dictionary

| Themes | Dictionary (Root Words) |
|---|---|
| Sorry and Shame | "미안" (sorry, apologetic)<br>"죄송" (sorry, apologize)<br>"용서" (forgiveness, forgive)<br>"잘못" (mistake, fault, wrongdoing)<br>"후회" (regret, remorse)<br>"죄" (sin, crime, guilt)<br>"책임" (responsibility, accountability)<br>"민폐" (nuisance, inconvenience)<br>"반성" (self-reflection, remorse) |
| Love and Gratitude | "사랑" (love)<br>"고맙" (thankful, grateful)<br>"감사" (gratitude, thanks)<br>"고생" (hardship, toil)<br>"수고" (effort, hard work, trouble)<br>"보고싶" (miss, long for)<br>"애틋" (tender, bittersweet, fond)<br>"소중" (precious, valuable)<br>"그리" (long for, yearn, miss) |
| Burdensomeness | "버겁" (overwhelming, too much to handle)<br>"부담" (burden, pressure)<br>"짐" (load, burden)<br>"감당" (handle, cope with, bear)<br>"힘들" (hard, difficult, tough)<br>"무겁" (heavy)<br>"지치" (exhausted, tired, worn out)<br>"헷갈" (confuse, mix up) |
| Despair | "포기" (give up, abandon)<br>"좌절" (frustration, despair)<br>"죽" (die, death)<br>"끝" (end, finish)<br>"아무것" (anything, nothing)<br>"헛되" (vain, futile)<br>"무의미" (meaningless)<br>"잊히" (be forgotten)<br>"없어지" (disappear, vanish, be gone)<br>"그만두" (quit, stop) |
| Post-mortem Affairs | "부탁" (request, favor)<br>"정리" (organize, tidy up, settle)<br>"남기" (leave, leave behind)<br>"처리" (handle, deal with, process)<br>"보험" (insurance)<br>"은행" (bank)<br>"장례" (funeral)<br>"통장" (bankbook, passbook)<br>"유서" (suicide note, last letter) |

**Supplementary Data Table 3: PCA-reduced Clusters labelled based on their centroid values**

| Theme-based PCA-reduced K-means Clusters | Description |
|---|---|
| 1. Love and gratitude | Dominant cluster centroids are love and gratitude across all 3 sections |
| 2. Sorry and shame (First and Last Section) | Dominant cluster centroids were sorry and shame in the introduction and conclusion |
| 3. Sorry and shame (First and Middle Section) | High cluster centroids were observed in the introduction and body for sorry and shame |
| 4. Sorry and Shame + Love and Gratitude | Dominant cluster centroids were sorry and shame and love and gratitude across all sections |
| 5. Theme-absent | No dominant cluster centroids |
| 6. Sorry and shame (Middle and Last Section) | Dominant cluster centroids were sorry and shame in the body and conclusion |
| 7. Sorry and shame (First section) | Dominant cluster centroid in the intro for sorry and shame |
| 8. Pervasive sorry and shame | Dominant cluster centroid across all sections for sorry and shame |
| 9. Sorry and shame (Last section) | Dominant cluster centroid in conclusion for sorry and shame |
| 10. Pervasive love and gratitude + sorry and shame (last section) | Dominant cluster centroid in all sections for love and gratitude and dominant cluster centroid in conclusion for sorry and shame |
| 11. Pervasive love and gratitude + sorry and shame (First and middle section) | Dominant cluster centroid in all sections for love and gratitude and dominant cluster centroid in intro and body for sorry and shame |
| 12. Sorry and shame (middle section) | Dominant cluster centroid in the body for sorry and shame |
| 13. Sorry and shame + Love and gratitude (First and last section) | Dominant cluster centroids in the intro and conclusion for sorry and shame and love and gratitude |
| 14. Pervasive sorry and shame + Post-mortem affairs (Middle and last section) | Dominant cluster centroids in all sections for sorry and shame and dominant cluster centroids in body and conclusion for post-mortem affairs |
| 15. Sorry and shame + Love and gratitude (First section) + Post-mortem affairs (Last section) | Relatively high cluster centroids for sorry and shame and love and gratitude in the intro and dominant cluster centroids in the last section for post-mortem affairs |
| **Sentiment-based PCA-reduced K-means Clusters** | **Description** |
| 1. Pervasive Hostility | High centroids scores of anger hatred and resentment |
| 2. Defeat + Exhaustion | High centroid scores of defeat (in body) and exhaustion (in conclusion) |
| 3. Hatred + Resentment | High centroids scores of hatred (in body) and resentment (in conclusion) |
| 4. Emotionally Flat | No dominant centroid score |
| 5. Affection + Happiness | High centroid scores of affection (in body) and happiness (in conclusion) |
| 6. Hopelessness + Affiliative Remarks | Dominant centroid scores of despair (in intro), defeat (in body and exhaustion (in conclusion) alongside affection (in body and happiness (in conclusion) |
| 7. Hopelessness + Hostility + Affiliative Remarks | Dominant centroid scores of defeat, hatred and affection (in body) and exhaustion, resentment and happiness (in conclusion) |
| 8. Emotional Turmoil | High centroid scores of despair, anger, defeat, hatred, affection, despair, exhaustion, resentment and happiness |
| 9. Defeat + Exhaustion + Happiness | Dominant centroid scores of defeat and affection (in body) and exhaustion and happiness (in conclusion) |
| 10. Despair + Defeat + Exhaustion | Dominant centroid score of despair (in intro), defeat (in body) and exhaustion (in conclusion) |

**Supplementary Data Table 4: Delta Lift Values across Sections in Gender**

**Gender: Male – Female: Section 1 → Section 2 (cellwise ΔLR)**

| Section 1 theme | Sorry and Shame | Love and Gratitude | Burden | Despair | Post-mortem Affairs |
|---|---|---|---|---|---|
| **Sorry and Shame** | -0.03 | -0.10** | -0.03 | 0.05 | -0.02 |
| **Love and Gratitude** | -0.14*** | -0.23* | -0.04 | 0.14 | 0.05 |
| **Burden** | -0.06 | -0.09 | 0.10 | 0.12 | -0.10 |
| **Despair** | 0.05 | 0.02 | 0.02 | 0.31 | 0.03 |
| **Post-mortem Affairs** | -0.07 | 0.02 | -0.01 | 0.19 | -0.61** |

**Gender: Male – Female: Section 2 → Section 3 (cellwise ΔLR)**

| Section 2 theme | Sorry and Shame | Love and Gratitude | Burden | Despair | Post-mortem Affairs |
|---|---|---|---|---|---|
| **Sorry and Shame** | -0.02 | -0.01 | 0.02 | 0.09 | -0.00 |
| **Love and Gratitude** | -0.05 | -0.05 | -0.05 | 0.14 | 0.01 |
| **Burden** | -0.00 | 0.13 | 0.08 | -0.12 | 0.08 |
| **Despair** | 0.04 | 0.04 | 0.00 | 0.20 | 0.06 |
| **Post-mortem Affairs** | 0.06 | -0.09 | 0.09 | -0.12 | 0.02 |

Asterisks denote statistically significant ΔLift values based on permutation testing (*$p < 0.05$, **$p < 0.01$, ***$p < 0.001$).

**Supplementary Data Table 5: Delta Lift Values across Sections between under 18 and rest**

**Age ≤18 vs Rest: Section 1 → Section 2 (cellwise ΔLR)**

| Section 1 theme | Sorry and Shame | Love and Gratitude | Burden | Despair | Post-mortem Affairs |
|---|---|---|---|---|---|
| **Sorry and Shame** | 0.17* | -0.08 | -0.19 | -0.04 | 0.13 |
| **Love and Gratitude** | 0.07 | -0.32 | -0.36 | -0.10 | -1.01*** |
| **Burden** | -0.01 | -0.48 | -0.12 | -0.14 | -0.31* |
| **Despair** | -0.19 | 0.11 | -0.17 | -0.02 | -1.00* |
| **Post-mortem Affairs** | -0.21 | 0.84 | 0.57 | 0.03 | 4.54*** |

**Age ≤18 vs Rest: Section 2 → Section 3 (cellwise ΔLR)**

| Section 2 theme | Sorry and Shame | Love and Gratitude | Burden | Despair | Post-mortem Affairs |
|---|---|---|---|---|---|
| **Sorry and Shame** | 0.00 | 0.02 | -0.26 | -0.31 | 0.06 |
| **Love and Gratitude** | -0.10 | -0.26 | -0.09 | -0.05 | 0.04 |
| **Burden** | 0.10 | 0.00 | 0.20 | -0.04 | -0.69 |
| **Despair** | -0.31 | -0.33 | -0.51 | 0.05 | -0.29 |
| **Post-mortem Affairs** | -0.04 | 0.23 | -0.68 | 0.69 | -1.61*** |

Asterisks denote statistically significant ΔLift values based on permutation testing (*p < 0.05, **p < 0.01, ***p < 0.001).

**Supplementary Data Table 6: Delta Lift Values across Sections between aged 19-34 and rest**

**Age 19-34 vs Rest: Section 1 → Section 2 (cellwise ΔLR)**

| Section 1 theme | Sorry and Shame | Love and Gratitude | Burden | Despair | Post-mortem Affairs |
|---|---|---|---|---|---|
| **Sorry and Shame** | 0.04 | -0.01 | -0.01 | -0.05 | 0.06 |
| **Love and Gratitude** | 0.11* | -0.05 | -0.04 | -0.05 | -0.12 |
| **Burden** | -0.07 | 0.18 | -0.10 | -0.02 | -0.06 |
| **Despair** | -0.02 | 0.03 | -0.15 | -0.15 | 0.15 |
| **Post-mortem Affairs** | 0.06 | 0.00 | -0.18 | -0.13 | 0.31 |

**Age 19-34 vs Rest: Section 2 → Section 3 (cellwise ΔLR)**

| Section 2 theme | Sorry and Shame | Love and Gratitude | Burden | Despair | Post-mortem Affairs |
|---|---|---|---|---|---|
| **Sorry and Shame** | 0.02 | 0.00 | -0.01 | -0.02 | -0.03 |
| **Love and Gratitude** | 0.09 | -0.25*** | 0.06 | 0.10 | 0.01 |
| **Burden** | 0.03 | -0.10 | 0.00 | -0.02 | 0.06 |
| **Despair** | -0.00 | -0.15 | 0.13 | -0.24 | -0.04 |
| **Post-mortem Affairs** | 0.01 | 0.06 | 0.16 | -0.17 | 0.50* |

Asterisks denote statistically significant ΔLift values based on permutation testing (*$p < 0.05$, **$p < 0.01$, ***$p < 0.001$).

**Supplementary Data Table 7: Delta Lift Values across Sections between aged 35-49 and rest**

**Age 35-49 vs Rest: Section 1 → Section 2 (cellwise ΔLR)**

| Section 1 theme | Sorry and Shame | Love and Gratitude | Burden | Despair | Post-mortem Affairs |
|---|---|---|---|---|---|
| **Sorry and Shame** | -0.02*** | 0.02 | -0.02 | 0.04 | -0.06 |
| **Love and Gratitude** | 0.03 | 0.19* | -0.03 | -0.07 | -0.04 |
| **Burden** | -0.04 | 0.03 | -0.26*** | 0.29* | -0.04 |
| **Despair** | -0.07 | 0.02 | 0.07 | -0.16 | -0.22 |
| **Post-mortem Affairs** | -0.12 | -0.02 | 0.21 | -0.05 | -0.37 |

**Age 35-49 vs Rest: Section 2 → Section 3 (cellwise ΔLR)**

| Section 2 theme | Sorry and Shame | Love and Gratitude | Burden | Despair | Post-mortem Affairs |
|---|---|---|---|---|---|
| **Sorry and Shame** | -0.07*** | -0.04 | -0.04 | -0.00 | 0.01 |
| **Love and Gratitude** | 0.03 | 0.01 | 0.12 | -0.17 | -0.03 |
| **Burden** | -0.07* | 0.00 | -0.22 | 0.01 | -0.08 |
| **Despair** | -0.02 | -0.01 | 0.04 | 0.13 | -0.01 |
| **Post-mortem Affairs** | -0.05 | 0.08 | -0.09 | 0.01 | -0.21 |

Asterisks denote statistically significant ΔLift values based on permutation testing (*p < 0.05, **p < 0.01, ***p < 0.001).

**Supplementary Data Table 8: Delta Lift Values across Sections between aged 50-64 and rest**

**Age 50-64 vs Rest: Section 1 → Section 2 (cellwise ΔLR)**

| Section 1 theme | Sorry and Shame | Love and Gratitude | Burden | Despair | Post-mortem Affairs |
|---|---|---|---|---|---|
| **Sorry and Shame** | -0.05** | 0.01 | -0.07 | -0.01 | -0.03 |
| **Love and Gratitude** | -0.05* | -0.06 | 0.04 | 0.07 | 0.02 |
| **Burden** | -0.05 | -0.16 | -0.02 | -0.32* | 0.01 |
| **Despair** | 0.07 | -0.08 | 0.08 | 0.18 | 0.08 |
| **Post-mortem Affairs** | 0.06 | -0.08 | 0.21 | 0.06 | -0.15 |

**Age 50-64 vs Rest: Section 2 → Section 3 (cellwise ΔLR)**

| Section 2 theme | Sorry and Shame | Love and Gratitude | Burden | Despair | Post-mortem Affairs |
|---|---|---|---|---|---|
| **Sorry and Shame** | -0.02 | -0.06 | 0.01 | -0.00 | 0.07 |
| **Love and Gratitude** | -0.06 | 0.08 | -0.17 | 0.04 | -0.09 |
| **Burden** | -0.10* | -0.18* | -0.13 | -0.03 | 0.21* |
| **Despair** | 0.04 | 0.15 | -0.15 | -0.27 | 0.08 |
| **Post-mortem Affairs** | 0.01 | -0.09 | 0.12 | 0.18 | -0.11 |

Asterisks denote statistically significant ΔLift values based on permutation testing (*p < 0.05, **p < 0.01, ***p < 0.001).

**Supplementary Data Table 9: Delta Lift Values across Sections between aged 65 and above and rest**

**Age 65+ vs Rest: Section 1 → Section 2 (cellwise ΔLR)**

| Section 1 theme | Sorry and Shame | Love and Gratitude | Burden | Despair | Post-mortem Affairs |
|---|---|---|---|---|---|
| **Sorry and Shame** | $0.05^{*}$ | 0.02 | $0.11^{*}$ | 0.01 | $0.19^{*}$ |
| **Love and Gratitude** | -0.03 | -0.09 | 0.19 | 0.04 | 0.10 |
| **Burden** | $0.18^{**}$ | -0.06 | $0.65^{***}$ | -0.22 | $0.43^{**}$ |
| **Despair** | 0.04 | -0.12 | -0.15 | -0.30 | $0.33^{*}$ |
| **Post-mortem Affairs** | 0.03 | 0.17 | $-0.57^{**}$ | 0.20 | 0.13 |

**Age 65+ vs Rest: Section 2 → Section 3 (cellwise ΔLR)**

| Section 2 theme | Sorry and Shame | Love and Gratitude | Burden | Despair | Post-mortem Affairs |
|---|---|---|---|---|---|
| **Sorry and Shame** | $0.05^{*}$ | -0.02 | -0.10 | $0.14^{*}$ | -0.03 |
| **Love and Gratitude** | $-0.13^{***}$ | $0.15^{*}$ | -0.26 | -0.00 | $0.19^{*}$ |
| **Burden** | $0.14^{***}$ | 0.04 | -0.21 | -0.23 | -0.04 |
| **Despair** | -0.04 | -0.07 | -0.19 | 0.25 | 0.18 |
| **Post-mortem Affairs** | 0.05 | 0.17 | -0.02 | -0.01 | -0.03 |

Asterisks denote statistically significant ΔLift values based on permutation testing (*p < 0.05, **p < 0.01, ***p < 0.001).

**Supplementary Data Table 10:**

**Gender: Male – Female: Section 1 to Section 2 (opening to middle)**

| Origin sentiment | Despair | Defeat | Exhaustion | Anxious | Disappointment | Anger | Hatred | Resentment | Sadness | Sorrow | Gratitude | Affection | Happiness | Relief | Neutral |
|---|---|---|---|---|---|---|---|---|---|---|---|---|---|---|---|
| **Despair** | -0.12 | -0.30 | -0.04 | 0.10 | -0.01 | 0.01 | -0.16 | -0.15 | -0.01 | -0.10 | 0.08 | -0.43 | -0.15 | 0.06 | 0.23 |
| **Defeat** | -0.09 | -1.80 | 0.17 | 0.26 | -0.00 | 0.04 | 0.57 | 0.57 | -0.04 | 0.43 | -0.28 | -1.18 | -0.75* | -0.56* | 0.28 |
| **Exhaustion** | -0.03 | -0.09 | 0.03 | 0.06 | -0.03 | 0.12 | 0.20 | -0.20 | -0.03 | -0.09 | 0.02 | -0.21 | -0.06 | 0.01 | 0.26* |
| **Anxious** | -0.01 | 0.22 | -0.06 | 0.08 | -0.05 | 0.18 | 0.23 | -0.13 | -0.06* | 0.05 | 0.19 | -0.45 | -0.02 | 0.13 | 0.19 |
| **Disappointment** | -0.02 | -0.08 | -0.02 | -0.01 | -0.03 | 0.01 | -0.03 | -0.04 | -0.01 | 0.03 | -0.02 | 0.04 | -0.07 | -0.08 | -0.04 |
| **Anger** | -0.05 | 0.16 | -0.01 | 0.09 | 0.00 | 0.11 | -0.44 | -0.12 | 0.04 | 0.06 | -0.04 | -1.15* | -0.04 | 0.18 | -0.07 |
| **Hatred** | -0.01 | 0.03 | -0.08 | -0.14 | -0.17 | 0.32 | 0.54 | 0.24 | -0.06 | -0.03 | 0.45 | 0.00 | 0.50 | 0.51 | -0.17 |
| **Resentment** | 0.15 | -1.87* | -0.04 | 0.22 | 0.07 | 0.93 | 1.24 | -0.46 | 0.07 | 0.05 | -0.31 | 0.00 | -0.82 | -0.18 | -0.72 |
| **Sadness** | -0.05* | -0.03 | -0.03 | 0.00 | -0.03* | -0.04 | 0.01 | 0.05 | -0.04*** | -0.06 | 0.01 | -0.09 | -0.13 | 0.01 | 0.04 |
| **Sorrow** | -0.07 | -0.02 | 0.13 | 0.10 | -0.05 | 0.17 | 0.25 | 0.00 | -0.01 | 0.17 | -0.04 | -0.81 | -0.36 | 0.11 | 0.07 |
| **Gratitude** | 0.06 | 0.16 | -0.01 | 0.05 | 0.04 | 0.16 | 0.21 | 0.17 | -0.08 | 0.11 | -0.11 | 0.72 | 0.24 | 0.34 | 0.17 |
| **Affection** | 0.35 | -0.42 | 0.39 | 0.49 | 0.00 | -0.06 | -0.51 | 2.93 | -0.11 | 0.25 | 0.86 | 15.19 | 2.38 | 0.98 | -1.50 |
| **Happiness** | 0.03 | -0.22 | -0.06 | -0.23 | -0.01 | 0.18 | -0.20 | 0.56 | -0.09 | 0.08 | -0.05 | 4.61 | 2.11* | 1.09* | 0.12 |
| **Relief** | -0.08 | 0.04 | -0.03 | 0.01 | 0.05 | 0.25 | 0.02 | 0.26 | -0.10 | -0.11 | -0.04 | -0.17 | 0.69 | 0.28 | 0.32 |
| **Neutral** | 0.14 | -0.37 | 0.19 | 0.01 | 0.01 | -0.30 | -0.58 | 0.51 | 0.06 | 0.18 | -0.24 | -0.66 | 0.11 | -0.28 | 0.25 |

**Gender: Male – Female: Section 2 to Section 3 (middle to final)**

| Origin sentiment | Despair | Defeat | Exhaustion | Anxious | Disappointment | Anger | Hatred | Resentment | Sadness | Sorrow | Gratitude | Affection | Happiness | Relief | Neutral |
|---|---|---|---|---|---|---|---|---|---|---|---|---|---|---|---|
| **Despair** | 0.10 | 0.40 | 0.05 | -0.02 | 0.01 | 0.01 | -0.15 | 0.04 | 0.02 | 0.18 | 0.14 | 0.47 | 0.06 | 0.09 | -0.16 |
| **Defeat** | 0.32 | 2.19 | 0.12 | 0.20 | 0.08 | 0.20 | 0.25 | -0.26 | 0.14* | 0.36 | 0.09 | -0.00 | 0.46 | -0.64* | 0.22 |
| **Exhaustion** | -0.03 | 0.41 | 0.12* | -0.01 | -0.00 | -0.06 | -0.05 | -0.01 | 0.01 | 0.25* | 0.12 | 0.34 | 0.12 | 0.10 | 0.03 |
| **Anxious** | 0.17* | 0.05 | 0.16* | 0.13 | 0.01 | 0.03 | 0.01 | 0.33 | -0.00 | 0.33* | -0.03 | 0.31 | -0.12 | 0.01 | -0.03 |
| **Disappointment** | -0.01 | 0.04 | -0.02 | -0.02 | -0.01 | 0.01 | 0.04 | 0.02 | -0.00 | 0.01 | 0.04 | -0.09 | -0.04 | -0.05 | 0.08 |
| **Anger** | 0.04 | 0.39 | -0.10 | -0.08 | -0.06 | 0.08 | -0.51 | 0.49 | 0.02 | -0.00 | 0.26* | 0.70 | 0.12 | -0.08 | -0.05 |
| **Hatred** | 0.01 | 0.72 | -0.14 | -0.08 | -0.00 | -0.58 | -0.83 | -2.59 | 0.06 | -0.39 | 0.32 | 2.15 | 0.26 | -0.15 | 0.32 |
| **Resentment** | 0.74* | 0.85 | 0.34 | 0.13 | 0.24 | -0.62 | -1.95 | 1.88 | 0.29** | 0.40 | -0.07 | 0.99 | 0.61 | -0.16 | 0.11 |
| **Sadness** | -0.01 | 0.00 | -0.02 | -0.01 | -0.01 | 0.03 | 0.08 | 0.19 | -0.00 | -0.01 | -0.02 | -0.01 | 0.01 | -0.03 | 0.11 |
| **Sorrow** | 0.09 | 0.71 | 0.05 | 0.05 | 0.01 | 0.12 | -0.13 | -0.38 | 0.01 | 0.26 | 0.08 | 0.39 | 0.27 | 0.06 | -0.15 |
| **Gratitude** | 0.01 | -0.54 | 0.03 | 0.20* | 0.10 | 0.19 | 0.27 | 0.30 | 0.00 | -0.02 | -0.41 | 0.25 | -0.16 | -0.00 | 0.04 |
| **Affection** | -0.54 | -0.00 | -0.21 | -0.84* | -0.25 | 0.20 | 0.48 | -0.00 | -0.20 | -0.55 | 0.16 | 15.43 | 5.66* | -0.23 | 0.41 |
| **Happiness** | 0.09 | 0.42 | -0.06 | -0.09 | 0.02 | 0.27 | 0.07 | -0.05 | -0.06 | -0.02 | -0.36 | 2.16 | 0.29 | 0.12 | -0.57 |
| **Relief** | 0.13 | 0.02 | 0.13 | 0.24 | 0.05 | 0.13 | 0.20 | -0.27 | 0.05 | 0.29 | -0.33 | 1.05 | -0.70 | -0.25 | -0.34 |
| **Neutral** | 0.01 | -0.04 | 0.09 | -0.11 | 0.02 | -0.03 | -0.38 | -2.94* | 0.02 | -0.07 | 0.15 | -0.23 | 0.14 | 0.26 | -1.09 |

*Note. ΔLift = difference in normalized transition lift between the focal group and comparison group. *p < 0.05, **p < 0.01, ***p < 0.001 (cellwise permutation tests).*

**Supplementary Data Table 11:**

**Age ≤18 vs Rest: Section 1 to Section 2 (opening to middle)**

| Origin sentiment | Despair | Defeat | Exhaustion | Anxious | Disappointment | Anger | Hatred | Resentment | Sadness | Sorrow | Gratitude | Affection | Happiness | Relief | Neutral |
|---|---|---|---|---|---|---|---|---|---|---|---|---|---|---|---|
| **Despair** | -0.04 | 0.07 | -0.06 | -0.17 | 0.19* | -0.11 | -0.25 | 0.32 | -0.00 | 0.03 | -0.39 | -0.31 | -0.07 | -0.47 | -0.49 |
| **Defeat** | -0.42 | -0.16 | -0.39 | 0.11 | -0.11 | -1.16 | -2.17 | 3.39 | -0.03 | 0.13 | -0.49 | -0.41 | -0.43 | -0.53 | -0.22 |
| **Exhaustion** | -0.01 | 0.07 | -0.05 | -0.05 | 0.09 | -0.33 | -0.16 | 0.67 | -0.00 | 0.10 | -0.50 | -0.48 | -0.45 | -0.40 | 0.34 |
| **Anxious** | 0.11 | 0.81 | -0.16 | -0.16 | -0.00 | 0.05 | 0.29 | -0.57 | 0.03 | -0.07 | -0.29 | -0.41 | -0.62 | -0.43 | -0.77 |
| **Disappointment** | -0.08 | 0.11 | -0.07 | -0.04 | 0.11* | -0.04 | -0.28 | 0.03 | 0.04 | -0.07 | -0.26 | -0.86 | -0.18 | -0.17 | 0.12 |
| **Anger** | -0.63 | -0.61 | -0.17 | -0.01 | 0.14 | -1.08 | -0.16 | -0.05 | -0.12 | -0.45 | -0.26 | -0.45 | 0.51 | 0.39 | -1.06 |
| **Hatred** | -0.55 | -1.11 | 0.22 | 0.43 | 0.15 | -1.11 | -2.33 | -1.11 | -0.04 | -0.76 | 0.16 | -0.00 | 0.88 | -0.57 | -0.87 |
| **Resentment** | -0.21 | -1.27 | 0.04 | 0.40 | 0.34 | -0.12 | 0.87 | -1.68 | 0.10 | -0.68 | -0.42 | -0.00 | -0.51 | -0.95 | -0.66 |
| **Sadness** | -0.00 | 0.28 | -0.07 | -0.08 | 0.07 | -0.22 | -0.07 | -0.00 | -0.02 | 0.04 | -0.05 | -0.92 | 0.15 | 0.21 | -0.62* |
| **Sorrow** | 0.38 | -0.09 | -0.02 | -0.31 | 0.07 | -0.08 | 0.15 | -0.34 | 0.05 | 0.40 | -0.44 | -0.48 | -0.50 | -0.58 | -0.24 |
| **Gratitude** | -0.19 | -0.42 | 0.10 | -0.36 | -0.32 | -0.26 | 0.20 | -0.28 | 0.09 | 0.07 | -0.00 | 6.18 | 0.47 | -0.06 | 1.94* |
| **Affection** | -1.05 | -1.38 | -0.84 | -0.57 | -0.98 | -0.39 | -0.28 | -1.71 | -0.23 | -0.89 | 3.25 | -8.35 | -3.44 | 6.89 | -0.54 |
| **Happiness** | -0.18 | -0.55 | 0.38 | -0.25 | -0.18 | -0.60 | -0.63 | -0.38 | 0.11 | 0.32 | 0.24 | 9.67 | -1.10 | -0.53 | 3.80* |
| **Relief** | 0.08 | -0.56 | 0.24 | -0.63 | -0.34 | -0.08 | 0.98 | -0.46 | 0.07 | 0.42 | 1.67 | -1.70 | 0.58 | 1.28 | -1.07 |
| **Neutral** | 1.05* | 2.15* | 0.19 | 0.26 | -0.01 | 2.35* | 2.49 | -1.09 | 0.10 | 0.92 | -0.67 | -0.73 | -0.65 | 0.89 | -5.60 |

**Age ≤18 vs Rest: Section 2 to Section 3 (middle to final)**

| Origin sentiment | Despair | Defeat | Exhaustion | Anxious | Disappointment | Anger | Hatred | Resentment | Sadness | Sorrow | Gratitude | Affection | Happiness | Relief | Neutral |
|---|---|---|---|---|---|---|---|---|---|---|---|---|---|---|---|
| **Despair** | 0.08 | -0.55 | -0.05 | -0.20 | -0.08 | -0.43 | -0.46 | -0.57 | -0.08 | 0.27 | -0.03 | -0.33 | 0.25 | 0.30 | 0.71 |
| **Defeat** | 0.51 | -0.26 | 0.53 | -0.43 | 0.32 | -0.53 | 0.14 | -0.43 | -0.15 | 1.60 | -0.51 | -0.00 | 0.15 | 0.13 | -0.40 |
| **Exhaustion** | 0.08 | -0.56 | -0.08 | -0.23 | 0.04 | -0.28 | -0.23 | -0.79 | -0.04 | -0.06 | 0.04 | -0.46 | 0.23 | 0.23 | 0.25 |
| **Anxious** | 0.16 | -0.69 | 0.19 | 0.23 | -0.08 | 0.05 | -0.48 | -0.72 | -0.02 | -0.07 | -0.14 | -0.80 | -0.10 | 0.20 | -0.33 |
| **Disappointment** | 0.11 | 0.05 | 0.08 | -0.29** | 0.09 | 0.05 | -0.18 | -0.71 | -0.04 | 0.33 | 0.11 | -0.09 | 0.20 | 0.06 | 0.55* |
| **Anger** | -0.03 | 1.10 | 0.11 | 0.79* | -0.11 | 0.17 | -0.45 | -2.64 | -0.01 | -0.00 | 0.21 | -0.47 | -0.13 | 0.17 | -0.88 |
| **Hatred** | -0.50 | -1.16 | -0.14 | 0.63 | 0.02 | 1.95 | 0.50 | -2.46 | 0.23 | 0.07 | -0.16 | -1.35 | -0.51 | 0.51 | -0.65 |
| **Resentment** | 0.26 | 1.37 | 0.09 | -0.20 | -0.26 | 1.15 | 2.74 | 12.54 | 0.03 | 1.53 | 0.12 | -0.68* | -0.41 | 0.93 | -1.49 |
| **Sadness** | -0.00 | -0.16 | -0.06 | -0.16 | -0.01 | -0.16 | -0.12 | -0.66 | -0.06 | -0.06 | 0.08 | 0.23 | 0.24 | 0.05 | 0.34 |
| **Sorrow** | 0.20 | -0.32 | -0.08 | -0.05 | -0.10 | 0.18 | 0.42 | -0.49 | -0.12 | 0.62 | -0.18 | -0.27 | 0.14 | 0.04 | -0.29 |
| **Gratitude** | -0.50 | -0.58 | -0.43 | 0.26 | -0.18 | -0.07 | 0.14 | -0.65 | -0.05 | -0.57 | -0.31 | 1.91 | 0.32 | -0.71 | 0.70 |
| **Affection** | -0.46 | -0.00 | -0.57 | -0.61 | -0.57 | -0.13 | -0.32 | -0.00 | 0.93 | -0.53 | -1.85 | -10.04 | -4.54 | -1.08 | -0.30 |
| **Happiness** | -0.54 | -0.59 | -0.41 | -0.18 | 0.17 | 0.07 | -0.19 | 7.80* | 0.03 | -0.40 | -0.51 | -0.97 | -1.47 | -0.21 | 1.82 |
| **Relief** | -0.14 | 0.60 | -0.19 | -0.00 | 0.05 | -0.60 | -0.40 | -0.73 | 0.33 | -0.09 | 0.11 | -2.00 | -0.60 | -0.18 | 3.23* |
| **Neutral** | -0.57 | -0.31 | 0.59 | -1.06 | -0.61 | -1.17 | -0.99 | -1.87 | -0.56 | -0.25 | 0.99 | -0.98 | -0.69 | -0.89 | -3.83 |

*Note. ΔLift = difference in normalized transition lift between the focal group and comparison group. *p < 0.05, **p < 0.01, ***p < 0.001 (cellwise permutation tests).*

**Supplementary Data Table 12:**

**Age 19-34 vs Rest: Section 1 to Section 2 (opening to middle)**

| Origin sentiment | Despair | Defeat | Exhaustion | Anxious | Disappointment | Anger | Hatred | Resentment | Sadness | Sorrow | Gratitude | Affection | Happiness | Relief | Neutral |
|---|---|---|---|---|---|---|---|---|---|---|---|---|---|---|---|
| **Despair** | -0.07 | -0.40 | -0.10 | -0.05 | -0.02 | 0.22$^{*}$ | 0.32 | -0.10 | -0.00 | -0.17 | -0.03 | 0.13 | 0.11 | -0.10 | -0.18 |
| **Defeat** | -0.36 | -1.70 | -0.29 | -0.04 | -0.13 | 0.17 | -0.48 | -0.60 | -0.04 | -0.98 | 0.47 | -0.65 | -0.22 | 0.46 | -0.30 |
| **Exhaustion** | -0.05 | -0.26 | -0.13$^{*}$ | -0.07 | -0.01 | 0.20$^{*}$ | 0.20 | 0.18 | -0.01 | -0.22 | -0.04 | -0.16 | -0.06 | -0.00 | -0.22 |
| **Anxious** | -0.06 | -0.10 | -0.08 | -0.01 | -0.06 | 0.10 | 0.08 | 0.23 | -0.03 | -0.04 | -0.15 | -0.10 | 0.12 | -0.00 | -0.21 |
| **Disappointment** | -0.05 | 0.10 | -0.01 | -0.02 | -0.02 | -0.05 | -0.03 | -0.15 | -0.01 | -0.01 | -0.05 | -0.00 | -0.05 | -0.02 | -0.32$^{***}$ |
| **Anger** | -0.08 | -0.06 | -0.07 | -0.08 | -0.02 | 0.29 | 0.22 | -0.91 | -0.01 | -0.05 | -0.01 | 1.30 | 0.08 | -0.17 | 0.53 |
| **Hatred** | -0.03 | 0.36 | 0.19 | -0.04 | -0.09 | 0.92 | 2.12 | 0.58 | -0.03 | 0.10 | -0.03 | -0.00 | -0.03 | -0.38 | -0.93 |
| **Resentment** | 0.14 | -0.32 | -0.02 | -0.25 | -0.30 | -0.72 | -0.24 | -0.32 | -0.02 | -0.09 | 0.62 | -0.00 | 0.76 | 0.91 | 0.83 |
| **Sadness** | -0.02 | -0.05 | -0.02 | -0.00 | -0.01 | -0.06 | -0.02 | -0.12 | -0.00 | -0.04 | -0.03 | 0.07 | -0.03 | -0.01 | -0.25$^{***}$ |
| **Sorrow** | -0.21 | -0.89 | -0.15 | -0.04 | -0.07 | 0.13 | 0.33 | -0.00 | -0.07 | -0.68$^{**}$ | 0.22 | -0.11 | 0.37 | -0.05 | -0.03 |
| **Gratitude** | -0.02 | -0.08 | -0.18 | -0.17 | -0.00 | -0.25 | -0.13 | -0.31 | -0.04 | -0.01 | -0.62 | -0.79 | -0.65 | 0.60 | 0.32 |
| **Affection** | 0.84 | 0.66 | 0.65 | 0.71 | -0.08 | -0.41 | -0.30 | -1.73 | -0.07 | -0.01 | -0.06 | 18.78 | -0.07 | -1.02 | -0.52 |
| **Happiness** | -0.01 | 0.49 | -0.20 | 0.19 | -0.15 | -0.67$^{*}$ | -0.73 | -0.42 | -0.17 | -0.04 | -0.50 | -4.94 | -0.95 | 0.30 | 0.20 |
| **Relief** | -0.14 | -0.39 | -0.12 | -0.21 | -0.00 | -0.59$^{**}$ | -0.37 | 0.23 | -0.07 | -0.13 | -0.43 | -1.98 | -0.32 | 0.14 | 2.33$^{***}$ |
| **Neutral** | -0.08 | -0.03 | 0.24 | 0.16 | -0.05 | -0.65 | -0.51 | -1.18 | -0.02 | -0.23 | 0.66$^{*}$ | -0.85 | -0.02 | 0.58 | 4.49$^{***}$ |

**Age 19-34 vs Rest: Section 2 to Section 3 (middle to final)**

| Origin sentiment | Despair | Defeat | Exhaustion | Anxious | Disappointment | Anger | Hatred | Resentment | Sadness | Sorrow | Gratitude | Affection | Happiness | Relief | Neutral |
|---|---|---|---|---|---|---|---|---|---|---|---|---|---|---|---|
| **Despair** | -0.15 | -0.47 | -0.15$^{*}$ | -0.07 | -0.02 | 0.04 | 0.04 | -0.02 | -0.07$^{*}$ | -0.25 | -0.06 | -0.42 | -0.23 | -0.03 | -0.12 |
| **Defeat** | -0.81$^{*}$ | -3.70$^{*}$ | -0.41 | 0.22 | -0.05 | -0.06 | -0.07 | 0.34 | -0.07 | -1.78$^{**}$ | 0.10 | -0.00 | 0.21 | -0.23 | -0.27 |
| **Exhaustion** | -0.06 | -0.36 | -0.13$^{*}$ | -0.00 | 0.04 | 0.08 | -0.08 | 0.02 | -0.05 | -0.13 | -0.16 | -0.39 | -0.21 | -0.07 | -0.07 |
| **Anxious** | -0.01 | -0.14 | -0.00 | -0.01 | -0.00 | 0.11 | 0.32 | 0.32 | -0.05 | -0.04 | -0.17 | -0.32 | -0.47$^{*}$ | -0.08 | 0.47$^{**}$ |
| **Disappointment** | -0.01 | -0.10 | -0.03 | -0.03 | -0.01 | -0.01 | -0.09 | -0.24 | -0.03$^{*}$ | -0.02 | -0.01 | -0.27 | 0.05 | -0.14$^{*}$ | -0.03 |
| **Anger** | -0.06 | -0.48 | 0.16 | 0.09 | -0.04 | 1.03$^{***}$ | 2.04$^{***}$ | 0.45 | -0.04 | -0.12 | 0.02 | -0.56 | -0.20 | -0.03 | -0.46 |
| **Hatred** | -0.02 | -0.78 | 0.24 | 0.40$^{*}$ | 0.04 | 1.43$^{**}$ | 2.84$^{**}$ | 2.26 | -0.01 | 0.18 | -0.11 | -1.65 | -0.19 | -0.25 | -0.30 |
| **Resentment** | -0.37 | -1.25 | -0.27 | -0.01 | 0.04 | 0.14 | 2.41 | -2.26 | -0.09 | -0.40 | 0.12 | -0.79 | 0.07 | 0.38 | -1.56 |
| **Sadness** | -0.03 | -0.16 | -0.02 | -0.01 | -0.00 | -0.02 | -0.13 | -0.19 | -0.03$^{*}$ | -0.04 | -0.10 | -0.22 | -0.09 | -0.06 | -0.11 |
| **Sorrow** | -0.38$^{*}$ | -0.39 | -0.21 | -0.06 | -0.00 | 0.10 | -0.05 | 0.14 | -0.09$^{*}$ | -0.39 | -0.07 | -0.37 | -0.12 | -0.03 | -0.24 |
| **Gratitude** | -0.06 | 0.27 | -0.14 | -0.22 | -0.04 | -0.31 | -0.21 | -0.24 | 0.12$^{*}$ | -0.10 | -0.25 | 0.16 | -0.37 | 0.72$^{*}$ | -0.00 |
| **Affection** | 0.59 | -0.00 | 0.36 | -0.35 | -0.25 | -0.16 | -0.38 | -0.00 | -0.27 | 0.92 | 0.41 | 15.35 | 2.96 | 0.03 | -0.34 |
| **Happiness** | -0.17 | 0.06 | -0.10 | -0.08 | -0.11 | -0.34 | -0.23 | 0.14 | 0.10 | -0.01 | -0.50 | 0.51 | -0.05 | 0.38 | 0.09 |
| **Relief** | 0.15 | 0.84 | -0.03 | -0.13 | -0.01 | -0.01 | -0.05 | 0.15 | 0.02 | -0.15 | 0.22 | 3.05$^{*}$ | 1.01 | 0.98$^{*}$ | -0.33 |
| **Neutral** | -0.08 | 0.50 | -0.08 | -0.04 | -0.34$^{***}$ | 0.33 | 0.22 | 1.08 | -0.20$^{*}$ | -0.08 | 0.24 | -1.06 | -0.29 | -0.12 | 2.47$^{**}$ |

*Note. ΔLift = difference in normalized transition lift between the focal group and comparison group. *p < 0.05, **p < 0.01, ***p < 0.001 (cellwise permutation tests).*

**Supplementary Data Table 13:**

**Age 35-49 vs Rest: Section 1 to Section 2 (opening to middle)**

| Origin sentiment | Despair | Defeat | Exhaustion | Anxious | Disappointment | Anger | Hatred | Resentment | Sadness | Sorrow | Gratitude | Affection | Happiness | Relief | Neutral |
|---|---|---|---|---|---|---|---|---|---|---|---|---|---|---|---|
| **Despair** | -0.08 | -0.01 | -0.04 | -0.12 | -0.01 | -0.24** | -0.40* | -0.14 | -0.02 | -0.02 | -0.07 | 0.35 | -0.13 | 0.03 | -0.03 |
| **Defeat** | -0.18 | -2.48* | -0.25 | -0.30 | -0.04 | -0.03 | 0.27 | 0.24 | -0.20*** | -0.97* | -0.40 | 1.39 | -0.15 | 0.20 | -0.31 |
| **Exhaustion** | -0.04 | 0.07 | -0.02 | -0.05 | -0.04 | -0.14 | -0.34* | -0.14 | -0.03 | -0.04 | -0.09 | 0.03 | -0.11 | -0.10 | -0.13 |
| **Anxious** | -0.01 | 0.12 | 0.03 | 0.07 | 0.04 | 0.18 | 0.05 | 0.45 | -0.01 | -0.08 | -0.13 | 0.20 | -0.12 | -0.21 | 0.07 |
| **Disappointment** | -0.01 | -0.02 | -0.01 | -0.04 | -0.00 | -0.07 | -0.02 | -0.01 | -0.01 | -0.01 | -0.07 | 0.14 | -0.03 | -0.10 | -0.01 |
| **Anger** | -0.04 | -0.07 | -0.02 | 0.08 | 0.04 | 0.46 | 1.09* | 1.26* | 0.02 | -0.24 | -0.07 | -0.58 | -0.14 | 0.08 | -0.20 |
| **Hatred** | 0.08 | 0.66 | -0.16 | -0.07 | 0.10 | 0.91 | 2.32* | 3.42** | -0.04 | -0.14 | -0.14 | -0.00 | -0.18 | -0.01 | -0.24 |
| **Resentment** | -0.21 | 0.15 | -0.10 | 0.11 | 0.12 | 0.82 | 0.58 | 4.01 | 0.04 | -0.28 | -0.32 | -0.00 | -0.12 | -0.47 | -0.85 |
| **Sadness** | -0.03 | -0.07 | -0.03 | -0.05* | -0.02 | -0.06 | -0.14* | 0.04 | -0.01 | -0.03 | -0.00 | -0.03 | -0.05 | -0.03 | -0.01 |
| **Sorrow** | -0.19 | -0.05 | -0.07 | -0.11 | -0.10* | -0.10 | -0.19 | 0.23 | -0.08* | -0.19 | -0.14 | 1.00* | -0.05 | -0.01 | -0.19 |
| **Gratitude** | -0.09 | -0.21 | -0.05 | -0.11 | -0.01 | 0.06 | -0.19 | -0.16 | -0.06 | -0.02 | 0.30 | -1.03 | 0.38 | -0.17 | 0.07 |
| **Affection** | 0.41 | 0.57 | -0.01 | 0.03 | 0.15 | -0.21 | -0.44 | 2.12 | 0.17 | 0.56 | -0.03 | -2.47 | -0.36 | 0.10 | -0.80 |
| **Happiness** | -0.03 | -0.44 | -0.02 | -0.06 | 0.17 | 0.33 | 0.23 | 0.04 | 0.09 | -0.07 | -0.53 | -3.14 | -1.02 | -0.83 | 0.27 |
| **Relief** | -0.07 | -0.06 | -0.01 | 0.18 | -0.02 | -0.08 | -0.16 | -0.60 | -0.08 | -0.16 | -0.02 | -0.40 | -0.97 | -0.29 | 0.22 |
| **Neutral** | -0.08 | 0.06 | -0.16 | 0.37* | -0.01 | 0.16 | 0.28 | -0.87 | -0.00 | 0.14 | -0.15 | 1.34 | 0.81* | -0.23 | 1.33 |

**Age 35-49 vs Rest: Section 2 to Section 3 (middle to final)**

| Origin sentiment | Despair | Defeat | Exhaustion | Anxious | Disappointment | Anger | Hatred | Resentment | Sadness | Sorrow | Gratitude | Affection | Happiness | Relief | Neutral |
|---|---|---|---|---|---|---|---|---|---|---|---|---|---|---|---|
| **Despair** | -0.16 | -0.57 | -0.11 | 0.12 | 0.02 | -0.11 | -0.11 | 0.18 | -0.07** | -0.35* | -0.11 | 0.03 | -0.17 | -0.07 | -0.04 |
| **Defeat** | 0.07 | 0.05 | -0.18 | -0.04 | 0.01 | 0.24 | 0.38 | 0.39 | -0.10 | 0.67 | -0.73** | 0.00 | -0.67 | 0.01 | 0.39 |
| **Exhaustion** | -0.18** | -0.39 | -0.13* | 0.01 | -0.03 | -0.11 | 0.02 | -0.26 | -0.06** | -0.34** | -0.12 | -0.07 | -0.17 | 0.05 | 0.02 |
| **Anxious** | -0.09 | -0.12 | -0.07 | 0.06 | -0.04 | -0.15 | -0.09 | -0.48 | 0.01 | -0.11 | 0.14 | -0.56 | 0.25 | 0.15 | -0.27 |
| **Disappointment** | -0.05 | 0.00 | -0.04 | 0.02 | -0.04** | -0.03 | -0.03 | -0.08 | -0.04** | -0.04 | 0.04 | -0.28 | -0.06 | 0.05 | -0.15* |
| **Anger** | 0.01 | 0.02 | 0.05 | -0.06 | 0.02 | 0.23 | 0.51 | -0.13 | -0.04 | 0.09 | -0.11 | 1.07* | -0.10 | 0.01 | 0.50* |
| **Hatred** | -0.30 | -0.59 | -0.09 | -0.04 | -0.14 | 0.79 | 0.58 | 0.85 | -0.17* | 0.03 | 0.02 | 3.12* | 0.27 | -0.10 | -0.03 |
| **Resentment** | 0.37 | 0.72 | 0.33 | 0.35 | -0.11 | 0.01 | -0.61 | 3.55 | -0.03 | 0.26 | 0.08 | 2.42* | 0.42 | -0.11 | -0.24 |
| **Sadness** | -0.06* | -0.11 | -0.07** | -0.05 | -0.03* | -0.05 | -0.04 | -0.13 | -0.04** | -0.10* | 0.03 | -0.12 | -0.10 | 0.01 | -0.19** |
| **Sorrow** | -0.27 | -0.87 | -0.25* | 0.02 | -0.08 | -0.08 | -0.12 | 0.14 | -0.14*** | -0.71** | -0.17 | 0.18 | -0.17 | 0.13 | 0.03 |
| **Gratitude** | -0.06 | -0.53 | -0.06 | -0.09 | 0.00 | 0.01 | -0.25 | 0.30 | -0.10* | -0.09 | -0.10 | -0.78 | 0.02 | -0.10 | -0.04 |
| **Affection** | 0.10 | 0.00 | 0.00 | 0.49 | 0.16 | -0.17 | -0.41 | 0.00 | 0.23 | -0.78 | 2.09* | -12.24 | 0.41 | -0.36 | -0.35 |
| **Happiness** | 0.04 | -0.03 | 0.00 | -0.07 | 0.05 | 0.16 | -0.05 | 0.46 | -0.07 | 0.02 | -0.07 | -0.43 | -0.15 | -0.14 | 0.52 |
| **Relief** | -0.10 | -0.56 | 0.01 | -0.04 | -0.04 | -0.03 | -0.25 | 0.79 | 0.00 | -0.20 | 0.16 | -2.42 | 0.36 | -0.12 | -0.16 |
| **Neutral** | -0.13 | 0.00 | -0.07 | 0.21 | -0.04 | 0.08 | 0.69 | 3.25** | 0.02 | -0.15 | -0.37 | 2.44* | 0.46 | 0.05 | 3.44*** |

*Note. ΔLift = difference in normalized transition lift between the focal group and comparison group. *p < 0.05, **p < 0.01, ***p < 0.001 (cellwise permutation tests).*

**Supplementary Data Table 14:**

**Age 50-64 vs Rest: Section 1 to Section 2 (opening to middle)**

| Origin sentiment | Despair | Defeat | Exhaustion | Anxious | Disappointment | Anger | Hatred | Resentment | Sadness | Sorrow | Gratitude | Affection | Happiness | Relief | Neutral |
|---|---|---|---|---|---|---|---|---|---|---|---|---|---|---|---|
| **Despair** | -0.04 | -0.14 | -0.04 | 0.07 | -0.03 | 0.06 | 0.10 | -0.02 | -0.07** | -0.30** | -0.05 | -0.42 | -0.02 | -0.06 | 0.06 |
| **Defeat** | -0.66** | -1.43 | -0.45** | 0.27 | -0.02 | -0.14 | 1.02 | -1.32 | -0.11 | 0.15 | -0.20 | -0.50 | 0.40 | -0.64* | 0.40 |
| **Exhaustion** | -0.04 | -0.23 | 0.05 | 0.01 | 0.01 | 0.05 | 0.15 | 0.05 | -0.03 | -0.05 | 0.09 | -0.04 | 0.09 | 0.02 | 0.07 |
| **Anxious** | 0.09 | -0.13 | 0.05 | -0.06 | 0.00 | -0.17 | -0.24 | -0.32 | 0.01 | 0.15 | 0.25* | 0.35 | 0.17 | 0.28* | 0.20 |
| **Disappointment** | -0.07* | -0.24** | -0.05* | 0.04 | -0.01 | 0.00 | -0.10 | 0.07 | -0.03** | -0.07 | 0.02 | -0.04 | -0.01 | 0.10 | 0.12 |
| **Anger** | 0.12 | 0.29 | -0.01 | 0.02 | -0.07 | -0.15 | -0.73 | 0.44 | -0.01 | 0.13 | 0.13 | 0.01 | 0.34 | 0.11 | -0.08 |
| **Hatred** | -0.24 | -1.33* | 0.01 | -0.04 | -0.11 | -0.53 | -1.81 | -1.72 | 0.00 | -0.07 | 0.12 | 0.00 | 0.18 | 0.41 | -0.08 |
| **Resentment** | -0.22 | 1.23 | 0.00 | 0.09 | 0.02 | 0.50 | -1.86 | -0.91 | -0.07 | -0.08 | -0.06 | 0.00 | -0.04 | 0.79 | -0.99 |
| **Sadness** | -0.06** | -0.17** | -0.02 | 0.02 | -0.01 | -0.02 | 0.04 | -0.06 | -0.03** | -0.08** | -0.02 | 0.05 | 0.01 | -0.02 | 0.10 |
| **Sorrow** | -0.16 | -1.02* | -0.19* | 0.05 | -0.01 | 0.14 | -0.03 | -0.11 | -0.06 | -0.28 | -0.22 | -0.62 | -0.27 | -0.06 | 0.18 |
| **Gratitude** | 0.03 | 0.40 | -0.02 | 0.08 | 0.07 | -0.06 | 0.18 | 0.46* | 0.06 | 0.04 | 0.26 | 0.92 | -0.12 | -0.38 | -0.20 |
| **Affection** | -0.08 | 0.49 | -0.28 | 0.05 | 0.10 | -0.56 | -0.41 | 0.15 | 0.04 | 0.08 | -0.74 | -2.76 | -3.13 | 0.02 | -0.83 |
| **Happiness** | 0.14 | -0.08 | -0.12 | -0.22 | 0.06 | -0.04 | 0.06 | 0.11 | -0.02 | 0.09 | 0.32 | 3.27 | 1.04 | -0.34 | -0.75 |
| **Relief** | 0.21 | 0.44 | 0.06 | -0.04 | 0.03 | 0.21 | 0.32 | 0.69 | 0.08 | 0.36 | 0.30 | -1.28 | 0.08 | -0.12 | -0.40 |
| **Neutral** | 0.12 | -0.21 | 0.10 | -0.05 | -0.04 | 0.54 | 0.54 | 0.90 | -0.05 | 0.12 | -0.44 | 0.73 | -0.57 | 0.13 | 0.14 |

**Age 50-64 vs Rest: Section 2 to Section 3 (middle to final)**

| Origin sentiment | Despair | Defeat | Exhaustion | Anxious | Disappointment | Anger | Hatred | Resentment | Sadness | Sorrow | Gratitude | Affection | Happiness | Relief | Neutral |
|---|---|---|---|---|---|---|---|---|---|---|---|---|---|---|---|
| **Despair** | -0.03 | 0.33 | 0.03 | -0.00 | 0.03 | 0.14 | 0.16 | 0.17 | -0.00 | -0.21 | -0.01 | 0.13 | 0.09 | -0.03 | 0.06 |
| **Defeat** | -0.15 | 1.64 | -0.02 | -0.08 | -0.12 | -0.07 | -0.16 | -0.53 | -0.06 | -0.98 | 0.45 | -0.00 | 0.28 | 0.22 | 0.13 |
| **Exhaustion** | -0.05 | 0.12 | 0.04 | -0.01 | -0.01 | 0.05 | 0.08 | 0.48 | -0.02 | -0.08 | 0.09 | 0.45 | 0.16 | -0.01 | 0.06 |
| **Anxious** | -0.01 | 0.05 | -0.00 | 0.02 | 0.05 | 0.17 | 0.10 | 0.16 | -0.02 | 0.08 | -0.10 | 0.51 | -0.01 | -0.11 | 0.38* |
| **Disappointment** | -0.05 | -0.15 | -0.01 | -0.00 | -0.01 | 0.02 | 0.12 | 0.19 | -0.00 | -0.14** | -0.04 | 0.18 | -0.09 | -0.00 | 0.07 |
| **Anger** | -0.01 | 0.29 | -0.32** | 0.06 | -0.01 | -0.44* | -0.86* | 0.26 | 0.06 | -0.12 | 0.14 | -0.09 | 0.42* | -0.13 | -0.05 |
| **Hatred** | 0.20 | 2.21* | -0.35* | -0.15 | 0.07 | -1.15** | -1.67 | -1.31 | 0.14* | -0.30 | 0.16 | -1.84 | 0.03 | -0.19 | 0.14 |
| **Resentment** | -0.18 | -1.94 | -0.07 | -0.27 | 0.08 | 0.14 | -0.45 | -4.04 | 0.17 | -0.21 | 0.10 | -0.91 | -0.05 | -0.18 | -0.27 |
| **Sadness** | -0.07* | 0.04 | -0.03 | -0.01 | -0.02 | 0.06 | 0.15* | 0.30* | -0.02* | -0.09* | -0.04 | 0.12 | -0.01 | -0.01 | 0.12 |
| **Sorrow** | 0.06 | -0.32 | 0.05 | 0.05 | -0.05 | 0.26 | 0.60* | 0.11 | -0.02 | -0.33 | -0.02 | -0.32 | 0.11 | -0.05 | 0.15 |
| **Gratitude** | -0.02 | 0.41 | 0.12 | 0.08 | -0.04 | 0.25 | 0.18 | 0.21 | -0.03 | 0.31* | -0.18 | 1.37 | -0.09 | -0.19 | -0.07 |
| **Affection** | -0.27 | -0.00 | -0.28 | 0.19 | 0.06 | -0.19 | -0.44 | -0.00 | -0.05 | 0.22 | -1.21 | 9.11 | 1.01 | -0.75 | -0.45 |
| **Happiness** | 0.08 | 0.24 | -0.06 | 0.08 | -0.01 | 0.14 | 0.23 | -0.43 | -0.06 | -0.04 | 0.11 | 1.98 | 0.55 | -0.19 | -0.28 |
| **Relief** | 0.15 | 0.37 | 0.17 | 0.12 | 0.04 | -0.03 | -0.21 | -0.64 | 0.06 | 0.51*** | -0.51* | 0.83 | -0.98* | -0.01 | -0.33 |
| **Neutral** | 0.39* | -0.38 | 0.21 | -0.01 | 0.14 | -0.41 | -0.88* | -1.45 | 0.17* | 0.34* | 0.29 | -1.41 | 0.15 | 0.09 | -0.76 |

*Note. ΔLift = difference in normalized transition lift between the focal group and comparison group. *p < 0.05, **p < 0.01, ***p < 0.001 (cellwise permutation tests).*

**Supplementary Data Table 15:**

**Age 65+ vs Rest: Section 1 to Section 2 (opening to middle)**

| Origin sentiment | Despair | Defeat | Exhaustion | Anxious | Disappointment | Anger | Hatred | Resentment | Sadness | Sorrow | Gratitude | Affection | Happiness | Relief | Neutral |
|---|---|---|---|---|---|---|---|---|---|---|---|---|---|---|---|
| **Despair** | -0.15* | -0.65** | -0.04 | -0.05 | -0.04 | 0.06 | -0.01 | 0.37 | -0.02 | -0.12 | 0.30** | 0.00 | 0.06 | 0.31** | 0.24 |
| **Defeat** | 1.07*** | 7.22*** | 1.04*** | -0.35 | 0.15 | 0.49 | -1.24 | 1.79* | 0.37*** | 0.97 | -0.07 | -0.46 | 0.66 | 0.14 | 0.74* |
| **Exhaustion** | -0.10 | -0.61** | -0.14** | 0.10 | 0.02 | -0.05 | 0.01 | -0.20 | -0.03 | -0.12 | 0.00 | 0.28 | 0.02 | 0.24* | 0.28* |
| **Anxious** | -0.17* | -0.49* | -0.05 | -0.03 | 0.01 | -0.11 | 0.15 | -0.22 | 0.00 | -0.13 | 0.07 | -0.52 | -0.08 | -0.05 | -0.13 |
| **Disappointment** | -0.05 | -0.05 | -0.01 | -0.03 | 0.00 | 0.06 | 0.19* | 0.05 | 0.00 | -0.01 | 0.04 | -0.03 | 0.04 | 0.04 | 0.00 |
| **Anger** | 0.12 | -0.46 | -0.03 | -0.08 | 0.04 | -0.55 | -0.82 | -1.48 | -0.00 | 0.38 | -0.03 | -0.57 | -0.61* | -0.14 | 0.16 |
| **Hatred** | 0.33 | 0.13 | -0.21 | -0.05 | 0.07 | -1.34* | -3.28* | -2.60 | 0.09 | 0.59 | -0.10 | 0.00 | -0.37 | -0.12 | 0.77 |
| **Resentment** | 0.60* | -1.33 | 0.15 | -0.10 | 0.10 | -1.04 | 2.03 | -3.14 | 0.06 | 1.42** | -0.15 | 0.00 | -0.59 | -0.94 | 1.42* |
| **Sadness** | 0.03 | -0.12 | 0.05* | 0.05* | 0.02 | 0.10* | 0.19* | 0.15 | 0.02 | 0.06 | -0.02 | 0.00 | -0.02 | 0.10 | 0.06 |
| **Sorrow** | 0.06 | -0.64 | -0.18 | -0.10 | 0.14** | -0.25 | -0.48 | -0.01 | 0.09* | -0.11 | 0.17 | -0.55 | -0.30 | 0.40* | 0.22 |
| **Gratitude** | 0.15 | 0.35 | 0.34*** | 0.34** | -0.02 | 0.28 | 0.10 | -0.08 | 0.04 | -0.04 | -0.19 | -0.10 | 0.01 | 0.15 | -0.01 |
| **Affection** | -0.84 | -1.48 | 0.41 | -0.36 | -0.21 | 1.21* | 1.48 | -2.17 | -0.20 | -0.97 | 0.71 | -0.23 | 6.24* | -0.58 | 1.46 |
| **Happiness** | -0.07 | 0.49 | 0.24 | 0.29 | -0.15 | 0.34 | 0.53 | 0.22 | 0.01 | -0.29 | 0.59 | 2.10 | 1.27 | 1.04 | 0.23 |
| **Relief** | 0.07 | 0.92* | 0.12 | 0.14 | 0.10 | 0.20 | -0.08 | -0.26 | 0.12 | 0.20 | -0.19 | 2.85 | 1.28* | -0.17 | -0.45 |
| **Neutral** | 0.05 | -0.23 | -0.03 | -0.36* | 0.13 | -0.48 | -0.63 | 0.42 | 0.14 | -0.04 | 0.22 | -1.12 | -0.09 | -0.39 | -3.09** |

**Age 65+ vs Rest: Section 2 to Section 3 (middle to final)**

| Origin sentiment | Despair | Defeat | Exhaustion | Anxious | Disappointment | Anger | Hatred | Resentment | Sadness | Sorrow | Gratitude | Affection | Happiness | Relief | Neutral |
|---|---|---|---|---|---|---|---|---|---|---|---|---|---|---|---|
| **Despair** | 0.02 | 0.96** | 0.06 | -0.08 | -0.07* | 0.10 | 0.07 | -0.45 | 0.06* | 0.98*** | 0.16 | 0.62* | 0.48** | 0.23* | 0.16 |
| **Defeat** | 0.09 | 0.25 | -0.08 | -0.17 | 0.00 | 0.19 | -0.47 | -0.48 | -0.08 | 0.58 | 0.65* | 0.00 | -0.47 | 0.39 | 0.27 |
| **Exhaustion** | 0.04 | 0.83** | 0.11 | 0.03 | -0.07* | 0.13 | 0.05 | -0.11 | 0.04 | 0.55*** | 0.12 | 0.22 | 0.13 | 0.05 | 0.15 |
| **Anxious** | 0.01 | 0.92** | -0.03 | -0.18* | -0.02 | -0.13 | -0.21 | 0.28 | 0.06* | -0.07 | 0.19 | 0.80 | 0.36* | 0.02 | -0.24 |
| **Disappointment** | 0.04 | 0.08 | 0.03 | 0.09* | 0.06*** | 0.03 | 0.02 | 0.13 | 0.05** | 0.07 | -0.11 | 0.57* | 0.08 | 0.10 | 0.13 |
| **Anger** | 0.19 | 0.32 | 0.20 | -0.17 | 0.08 | -0.60* | -1.19** | -0.30 | 0.04 | 0.38 | -0.15 | -0.55 | -0.18 | 0.15 | -0.10 |
| **Hatred** | 0.21 | 0.65 | 0.37* | -0.23 | 0.06 | -1.06* | -1.28 | -0.86 | -0.03 | -0.09 | -0.08 | 0.85 | -0.04 | 0.52* | 0.11 |
| **Resentment** | 0.23 | 3.88** | -0.09 | -0.05 | 0.10 | -0.53 | -1.10 | 0.71 | -0.05 | 0.47 | -0.52 | -0.82 | -0.46 | -0.10 | 0.80 |
| **Sadness** | 0.12** | 0.23* | 0.09** | 0.10** | 0.05** | 0.06 | 0.05 | 0.02 | 0.06*** | 0.15** | 0.00 | 0.23 | 0.14 | 0.08 | 0.18* |
| **Sorrow** | 0.09 | 0.14 | 0.20 | 0.01 | 0.11 | -0.39* | -0.69* | -0.55 | 0.19*** | 1.45*** | 0.27 | 1.39* | 0.23 | 0.09 | 0.29 |
| **Gratitude** | 0.36** | 0.22 | 0.21* | 0.27* | 0.14* | 0.02 | 0.29 | -0.32 | 0.02 | -0.14 | 0.65* | -2.29 | -0.02 | -0.11 | 0.12 |
| **Affection** | -0.54 | 0.00 | 0.03 | -0.43 | 0.04 | 0.49 | 1.17 | 0.00 | 0.06 | -0.60 | -1.10 | -12.44 | -5.21 | 1.12 | 0.66 |
| **Happiness** | 0.21 | -0.57 | 0.36* | 0.13 | 0.05 | -0.05 | 0.08 | -1.19 | 0.01 | -0.02 | 0.45 | -3.57 | -1.21 | 0.19 | -0.19 |
| **Relief** | -0.05 | -0.85 | -0.03 | -0.02 | 0.05 | 0.09 | 0.49* | -0.07 | -0.02 | -0.10 | 0.52 | -0.55 | 0.24 | -0.55 | 0.05 |
| **Neutral** | 0.00 | 0.58 | 0.00 | -0.16 | 0.06 | 0.18 | 0.17 | -1.54 | 0.04 | 0.04 | 0.15 | 0.04 | -0.07 | -0.19 | -2.80** |

*Note. ΔLift = difference in normalized transition lift between the focal group and comparison group. *p < 0.05, **p < 0.01, ***p < 0.001 (cellwise permutation tests).*
